# Predictions of arrhythmic, heart failure and mortality outcomes in pericarditis using automatic electrocardiogram analysis

**DOI:** 10.1101/2021.11.07.21266025

**Authors:** Ishan Lakhani, Jiandong Zhou, Michelle Vangi Wong, Sharen Lee, Jovy Ka Yiu Lau, Silverdew Shi, Wing Tak Wong, Tong Liu, Qingpeng Zhang, Gary Tse

## Abstract

**Background:** Pericarditis is a relatively rare disease with a global burden. Despite its strong association with adverse cardiovascular outcomes, identification of patients at risk of future heart failure or arrhythmic events is difficult. In the following study, automated electrocardiogram (ECG) were used to predict new onset ventricular tachycardia/fibrillation (VT/VF), atrial fibrillation (AF) and heart failure with reduced ejection fraction (HF) in an Asian cohort of pericarditis patients.

**Methods:** Consecutive patients admitted to a single tertiary center in Hong Kong, China, for a diagnosis of pericarditis between 1st January 2005 and 31st December 2019 with baseline ECG measurements were included. Patients with existing AF or HF were excluded. The follow-up period was until the 31st December 2020, or death. Cox regression was applied to identify significant predictors of the primary outcomes (incident VT/VF, AF or HF).

**Results:** A total of 874 patients were included. The cohort was 57% male and had a median age of 59 (IQR: 50-70) years old. During follow-up, 57 patients (6.5%), 156 (17.8%) and 168 (19.2%) suffered from VT/VF, AF and HF, respectively. Cox regression identified baseline VT/VF, terminal angle of the QRS vector in the transverse plane, mean QRS duration and mean QTc intervals as significant predictors of incident VT/VF events, with only the former most maintaining significance in multivariate analysis. In contrast, baseline age, prior diagnoses of hypertension, initial angle and magnitude of the QRS vector in the transverse plane, P-wave and QRS axis in the frontal plane, ST segment axis in the frontal and horizontal planes, mean PT interval, mean PR segment duration and QTc intervals were all univariate predictors of incident AF, albeit only baseline age and initial angel of the QRS vector in the transverse plane retained significance after multivariate adjustment. As it pertains to new-onset HF, several clinical and electrocardiographic parameters demonstrated an association with HF in univariate analysis, with prior diagnosis of HT or DM, initial QRS angle in transverse plane, I 40 in horizontal axis, ST-segment axis in the horizontal plane, T-wave frontal axis and atrial rate, of which, except for prior diagnosis of DM, I40 in horizontal axis and T-wave frontal axis, all variables showcased significant relationships in multivariate analysis

**Interpretation:** AF and HF are relatively common complications VT/VF occurs less frequently in the context of pericarditis. Different clinical and ECG predictors of these outcomes were identified. Future studies are still needed to evaluate their use for risk stratification in the clinical setting.

## Introduction

Acute pericarditis, an inflammation of the pericardial layer of the heart, is an immune-mediated clinical condition that is often unrecognized and associated with numerous of complications ^1^. Previous cohort studies have explored the aetiology ^2^, clinical presentation ^3, 4^ and risk factors leading to readmission ^5^, composite of cardiac tamponade, constrictive pericarditis, failed therapy, recurrence and mortality ^6^. By contrast, few studies have explored atrial or ventricular arrhythmic outcomes. Moreover, recent studies have reported the roles of automated electrocardiogram (ECG) analysis to facilitate risk stratification and prediction. However, few studies have examined the roles of ECG predictors for adverse outcomes in the pericarditis population. Therefore, the aim of this study is to investigate the predictive roles of automated electrocardiograms (ECG) for predicting new onset atrial fibrillation (AF), ventricular tachycardia/fibrillation (VT/VF) or heart failure (HF) in a cohort of pericarditis patients admitted to a single tertiary centre.

## Methods

### Study design and population

The study was approved by The Joint Chinese University of Hong Kong – New Territories East Cluster Clinical Research Ethics Committee. This was a retrospective, territory-wide cohort study of pericarditispatients with baseline ECG measurementsfrom 1^st^ January 2005to31^st^ December2019admitted to a single tertiary center fromthe Hong Kong region of China. The patients were identified from the Clinical Data Analysis and Reporting System (CDARS), a territory-wide database that centralizes patient information from 43 local hospitals and their associated ambulatory and outpatient facilities to establish comprehensive medical data, including clinical characteristics, disease diagnosis, laboratory results, and drug treatment details. The system has been previously used by both our team and other teams in Hong Kong^7, 8^. Patients’demographics, priorcomorbidities, medication prescriptions, laboratory examinations of complete blood counts, biochemical liver and renal tests, lipid and glucose tests and cardiac function tests, and ECG measurements were extracted.The list of ICD-9 codes for comorbidities and outcomes were detailed in the **Supplementary Table 1**.

### Baseline characteristics and ECG measurements

Clinical data was extracted from electronic health records. The following baseline clinical data were collected: 1) sex; 2) age of initial Brugada pattern presentation; 3) follow-up period; 4) type of Brugada pattern and presence of fever at initial presentation; 5) family history of BrS and VF/ SCD; 6) manifestation of syncope and if present, the number of episodes; 7) manifestation of VT/VF and if present, the number of episodes; 8) sodium channel blocker challenge test and results; 9) concomitant presence of other arrhythmia; 10) implantation of ICD. Patients presented with two or more episodes of VT/VF were defined to be of high VT/VF burden. Automatically measured parameters from ECG related to the P, Q, R, S and T-wave were extracted. This has been done previously by our team and further details could be found here ^9, 10^.

### Outcomes and statistical analysis

The primary outcome was new onset VT/VF. Secondary outcomes included new onset 1) AF, 2) new-onset HFrEF and 3) all-cause mortality. The secondary outcomes were cardiovascular mortality and all-cause mortality. Mortality data were obtained from the Hong Kong Death Registry, a population-based official government registry with the registered death records of all Hong Kong citizens linked to CDARS. There was no adjudication of the outcomes as this relied on the ICD-9 coding or a record in the death registry. However, the coding was performed by the clinicians or administrative staff, who were not involved in the mode development. Descriptive statistics are used to summarize baseline clinical characteristics of all pericarditis patients and based on the occurrence of the primary and secondary outcomes. Continuous variables were presented as median (95% confidence interval[CI] or interquartile range[IQR]) and categorical variables were presented as count (%). The Mann-Whitney U test was used to compare continuous variables. The χ2 test with Yates’ correction was used for 2×2 contingency data. Univariate logistic regression identifies significant mortality risk predictors. Hazard ratios (HRs) with corresponding 95% CIs and P-values were reported. There was no imputation performed for missing data. No blinding was performed for the predictor as the values were obtained from the electronic health records automatically.All statistical tests were two-tailed and considered significant if p value<0.001. They were performed using RStudio software (Version: 1.1.456) and Python (Version: 3.6).

## Results

### Basic characteristics

The baseline clinical characteristics of patients with/without spontaneous VT/VF are presented in **Table 1**. Subjects who developed incident VT/VF tended to have baseline VT/VF prior to the diagnosis of pericarditis. Regarding blood parameters, these individuals also reported significantly higher eosinophil counts, low-density-lipoprotein (LDL) levels, high-sensitivity troponin I (hsTnI) levels and lactate dehydrogenase (LDH) levels, as well as a lower mean corpuscular volume (MCV). Patients who developed VT/VF likewise had greater terminal QRS angles in the transverse plane and T-wave frontal axis, along with a longer QTc interval.

**Table 1.** Baseline clinical characteristics of patients with/without spontaneous VT/VF. * for p≤ 0.05, ** for p ≤ 0.01, *** for p ≤ 0.001. # indicates that the comparisons were made between patients with/without spontaneous VT/VF.

indicates that the comparisons were made between patients with/without spontaneous VT/VF.
| Characteristics | All patients (N=874)<br>Median (IQR);N or Count(%) | Spontaneous VT/VF (N=57)<br>Median (IQR);N or Count(%) | No spontaneous VT/VF (N=817)<br>Median (IQR);N or Count(%) | P value<sup>#</sup> |
| --- | --- | --- | --- | --- |
| ***Demographics*** |  |  |  |  |
| Male gender | 494(56.52%) | 30(52.63%) | 464(56.79%) | 0.8325 |
| Female gender | 380(43.47%) | 27(47.36%) | 353(43.20%) | 0.7972 |
| Baseline age, years | 59.43(49.5-69.76);n=874 | 58.84(47.25-68.06);n=57 | 59.51(49.82-69.86);n=817 | 0.477 |
| <40 | 119(13.61%) | 12(21.05%) | 107(13.09%) | 0.2144 |
| [40, 50) | 105(12.01%) | 6(10.52%) | 99(12.11%) | 0.9139 |
| [50-60) | 221(25.28%) | 13(22.80%) | 208(25.45%) | 0.8468 |
| [60-70) | 216(24.71%) | 14(24.56%) | 202(24.72%) | 0.8946 |
| [70-80) | 145(16.59%) | 10(17.54%) | 135(16.52%) | 0.9902 |
| $\geq 80$ | 68(7.78%) | 2(3.50%) | 66(8.07%) | 0.3607 |
| Charlson standard comorbidity index | 2.0(1.0-4.0);n=874 | 2.0(0.0-4.0);n=57 | 2.0(1.0-4.0);n=817 | 0.3003 |
| ***Past comorbidities*** |  |  |  |  |
| Baseline VT/VF | 8(0.91%) | 4(7.01%) | 4(0.48%) | <0.0001*** |
| Cerebrovascular disease | 28(3.20%) | 2(3.50%) | 26(3.18%) | 0.7969 |
| Rheumatic disease | 22(2.51%) | 4(7.01%) | 18(2.20%) | 0.0862 |
| Renal diseases | 48(5.49%) | 3(5.26%) | 45(5.50%) | 0.821 |
| Diabetes mellitus | 25(2.86%) | 3(5.26%) | 22(2.69%) | 0.4992 |
| Hypertension | 155(17.73%) | 11(19.29%) | 144(17.62%) | 0.9281 |
| AMI | 23(2.63%) | 0(0.00%) | 23(2.81%) | 0.405 |
| COPD | 8(0.91%) | 1(1.75%) | 7(0.85%) | 0.9702 |
| IHD | 86(9.83%) | 6(10.52%) | 80(9.79%) | 0.9477 |
| PVD | 12(1.37%) | 1(1.75%) | 11(1.34%) | 0.7372 |
| Stroke/TIA | 25(2.86%) | 2(3.50%) | 23(2.81%) | 0.9082 |
| Gastrointestinal bleeding | 39(4.46%) | 2(3.50%) | 37(4.52%) | 0.9892 |
| Immune mediated enterocolitis | 47(5.37%) | 2(3.50%) | 45(5.50%) | 0.7547 |
| Anxiety disorder and depression | 17(1.94%) | 1(1.75%) | 16(1.95%) | 0.6962 |
| SCD | 25(2.86%) | 1(1.75%) | 24(2.93%) | 0.9257 |
| Atrial flutter | 6(0.68%) | 0(0.00%) | 6(0.73%) | 0.853 |
| Accident fall | 50(5.72%) | 3(5.26%) | 47(5.75%) | 0.8812 |
| Malignancy | 165(18.87%) | 8(14.03%) | 157(19.21%) | 0.523 |
| Metastatic solid tumor | 94(10.75%) | 2(3.50%) | 92(11.26%) | 0.1429 |
| ***Medications*** |  |  |  |  |
| ACEI | 77(8.81%) | 7(12.28%) | 70(8.56%) | 0.5317 |
| ARB | 30(3.43%) | 5(8.77%) | 25(3.05%) | 0.0728 |
| Lipid-lowering drugs | - | - | - | - |
| Anticoagulants | 100(11.44%) | 11(19.29%) | 89(10.89%) | 0.145 |
| Insulin | - | - | - | - |
| Incretin mimetics | 10(1.14%) | 1(1.75%) | 9(1.10%) | 0.8411 |
| ***Complete blood counts*** |  |  |  |  |
| Mean corpuscular volume, fL | 89.8(86.3-93.35);n=543 | 88.5(85.0-90.6);n=37 | 90.0(86.4-93.5);n=506 | 0.0367* |
| Eosinophil, x10<sup>9</sup>/L | 0.1(0.0-0.2);n=504 | 0.2(0.1-0.3);n=33 | 0.1(0.0-0.2);n=471 | 0.0008*** |
| Lymphocyte, x10<sup>9</sup>/L | 1.15(0.8-1.7);n=505 | 1.1(0.8-1.6);n=33 | 1.15(0.8-1.7);n=472 | 0.9463 |
| Monocyte, x10<sup>9</sup>/L | 0.6(0.4-0.8);n=505 | 0.7(0.5-0.9);n=33 | 0.6(0.4-0.77);n=472 | 0.0509 |
| Neutrophil, x10<sup>9</sup>/L | 4.9(3.4-7.7);n=505 | 5.2(4.1-6.6);n=33 | 4.9(3.4-7.75);n=472 | 0.5897 |
| White cell count, x10<sup>9</sup>/L | 7.0(5.5-9.15);n=543 | 7.6(6.5-10.1);n=37 | 6.95(5.5-9.0);n=506 | 0.056 |
| Mean cell haemoglobin, pg | 30.6(29.0-32.0);n=543 | 29.7(28.5-31.3);n=37 | 30.7(29.1-32.1);n=506 | 0.0608 |
| Platelet, x10<sup>9</sup>/L | 212.0(166.5-275.0);n=543 | 243.0(197.0-294.0);n=37 | 209.0(165.0-274.0);n=506 | 0.0958 |
| Red blood count, x10<sup>12</sup>/L | 4.09(3.6-4.58);n=543 | 4.1(3.4-4.53);n=37 | 4.08(3.6-4.58);n=506 | 0.6528 |
| ***Liver and renal functions*** |  |  |  |  |
| K/Potassium, mmol/L | 4.0(3.7-4.4);n=684 | 4.0(3.6-4.35);n=52 | 4.0(3.7-4.4);n=632 | 0.5585 |
| Urate, mmol/L | 0.38(0.28-0.49);n=100 | 0.34(0.24-0.59);n=13 | 0.38(0.28-0.47);n=87 | 0.7701 |
| Albumin, g/L | 35.9(30.5-40.0);n=681 | 34.85(31.2-39.15);n=52 | 35.9(30.5-40.1);n=629 | 0.6382 |
| Na/Sodium, mmol/L | 138.45(135.2-140.9);n=684 | 138.8(136.05-140.75);n=52 | 138.4(135.05-140.9);n=632 | 0.7475 |
| Urea, mmol/L | 5.8(4.26-8.41);n=683 | 6.02(4.12-12.04);n=52 | 5.73(4.27-8.4);n=631 | 0.6471 |
| Protein, g/L | 71.0(65.6-75.4);n=681 | 71.25(67.05-75.35);n=52 | 71.0(65.5-75.4);n=629 | 0.9386 |
| Creatinine, umol/L | 84.0(66.0-113.0);n=682 | 89.0(59.5-122.0);n=51 | 83.0(66.0-111.5);n=631 | 0.7487 |
| Alkaline phosphatase, U/L | 82.0(65.0-118.0);n=681 | 85.0(69.0-118.5);n=52 | 82.0(64.0-118.0);n=629 | 0.4949 |
| Aspartate transaminase, U/L | 29.5(18.0-58.1);n=82 | 29.0(17.0-47.0);n=10 | 29.5(18.0-62.5);n=72 | 0.6967 |
| Alanine transaminase, U/L | 24.0(16.0-44.0);n=681 | 19.5(14.0-44.5);n=52 | 24.0(16.0-44.0);n=629 | 0.1168 |
| Bilirubin, umol/L | 10.4(6.9-16.8);n=681 | 8.35(6.0-13.0);n=52 | 10.6(7.0-17.0);n=629 | 0.0766 |
| ***Lipid and glucose tests*** |  |  |  |  |
| Triglyceride, mmol/L | 1.01(0.75-1.44);n=167 | 1.47(1.23-1.6);n=13 | 1.0(0.74-1.42);n=154 | 0.0864 |
| Low-density lipoprotein, mmol/L | 2.22(1.61-2.86);n=153 | 2.45(1.66-2.86);n=11 | 2.22(1.62-2.84);n=142 | 0.9859 |
| High-density lipoprotein, mmol/L | 1.07(0.81-1.41);n=153 | 0.86(0.78-1.0);n=11 | 1.12(0.82-1.48);n=142 | 0.0273* |
| Cholesterol, mmol/L | 3.82(3.15-4.72);n=167 | 4.05(3.42-4.82);n=13 | 3.82(3.15-4.71);n=154 | 0.9167 |
| HbA1c, g/dL | 12.3(10.8-13.7);n=543 | 11.4(10.6-13.7);n=37 | 12.3(10.8-13.7);n=506 | 0.3974 |
| Glucose, mmol/L | 6.41(5.44-8.16);n=467 | 6.65(5.73-7.9);n=36 | 6.4(5.43-8.2);n=431 | 0.5478 |
| ***Cardiac, clotting, inflammatory, and acid-base tests*** |  |  |  |  |
| D-dimer, ng/mL | 2666.0(1276.23-6132.04);n=73 | 2422.73(2332.06-3120.41);n=9 | 2684.52(1164.69-6470.84);n=64 | 0.9131 |
| High sensitive troponin-I, ng/L | 0.02(0.01-0.07);n=444 | 0.04(0.02-0.16);n=32 | 0.02(0.01-0.07);n=412 | 0.0155* |
| Lactate dehydrogenase, U/L | 265.0(211.0-387.5);n=483 | 304.5(233.0-440.5);n=36 | 262.0(207.0-371.5);n=447 | 0.0428* |
| APTT, second | 33.0(30.2-36.6);n=721 | 34.5(30.7-37.0);n=53 | 33.0(30.2-36.45);n=668 | 0.1871 |
| Prothrombin time/INR, second | 11.9(10.8-13.7);n=721 | 11.7(10.4-13.3);n=53 | 11.9(10.8-13.7);n=668 | 0.4269 |
| C-reactive protein, mg/dL | 2.91(0.56-8.62);n=556 | 3.76(0.73-7.04);n=40 | 2.78(0.56-8.67);n=516 | 0.6553 |
| ***ECG measurements*** |  |  |  |  |
| Trans QRS initangle | 42.0(19.0-90.0);n=574 | 38.5(14.0-94.0);n=36 | 42.0(19.0-90.0);n=538 | 0.5997 |
| Trans QRS initmag | 0.31(0.18-0.5);n=574 | 0.33(0.15-0.48);n=36 | 0.31(0.18-0.5);n=538 | 0.8278 |
| Trans QRS max angle | 331.0(42.5-347.0);n=574 | 323.0(76.0-348.0);n=36 | 331.0(42.5-347.0);n=538 | 0.5825 |
| Trans QRS max mag | 1.23(0.86-1.66);n=574 | 1.15(0.85-1.63);n=36 | 1.23(0.86-1.66);n=538 | 0.4962 |
| Trans QRS term angle | 243.0(214.0-273.0);n=574 | 225.5(201.0-248.5);n=36 | 244.0(215.5-273.0);n=538 | 0.0183* |
| Trans QRS term mag | 0.3(0.2-0.41);n=574 | 0.3(0.24-0.51);n=36 | 0.3(0.19-0.41);n=538 | 0.2255 |
| Trans QRS cwrot | -100.0(-100.0--87.5);n=574 | -100.0(-100.0--10.0);n=36 | -100.0(-100.0--100.0);n=538 | 0.3281 |
| P wave front axis | 54.0(39.0-68.0);n=798 | 54.5(40.0-69.5);n=54 | 54.0(39.0-68.0);n=744 | 0.6899 |
| P wave horizontal axis | 20.0(4.0-37.0);n=830 | 21.0(3.0-35.0);n=55 | 20.0(4.0-37.0);n=775 | 0.7535 |
| I 40 front axis | 38.0(15.0-58.0);n=860 | 34.0(8.5-53.0);n=55 | 38.0(15.0-58.0);n=805 | 0.177 |
| I 40 horizon axis | 28.0(11.5-44.0);n=839 | 28.0(17.0-48.5);n=55 | 28.0(11.0-44.0);n=784 | 0.4206 |
| QRS front axis | 52.0(20.0-75.0);n=871 | 48.0(10.0-79.0);n=57 | 52.0(21.0-75.0);n=814 | 0.7057 |
| QRS horizon axis | -11.0(-30.0-9.0);n=853 | -8.0(-46.5-20.0);n=56 | -11.0(-30.0-8.0);n=797 | 0.8747 |
| T wave 40ms front axis | 69.0(19.0-101.5);n=862 | 84.0(8.0-118.0);n=57 | 69.0(19.0-100.0);n=805 | 0.2919 |
| T wave 40ms horizon axis | -23.0(-52.0-184.0);n=829 | -10.5(-55.0-211.0);n=56 | -23.0(-52.0-179.0);n=773 | 0.5137 |
| ST segment front axis | 63.0(30.0-113.5);n=827 | 72.0(33.5-126.0);n=56 | 63.0(30.0-112.5);n=771 | 0.5575 |
| ST segment horizon axis | 76.0(50.0-118.0);n=780 | 79.0(51.0-143.5);n=51 | 76.0(50.0-117.0);n=729 | 0.4827 |
| T wave front axis | 45.0(14.0-72.0);n=857 | 50.0(35.0-101.0);n=55 | 44.0(13.0-70.5);n=802 | 0.0322* |
| T wave horizon axis | 46.0(25.0-76.0);n=805 | 48.0(16.0-84.0);n=53 | 46.0(25.0-75.5);n=752 | 0.8192 |
| Atrial rate | 80.0(67.0-99.5);n=874 | 83.0(75.0-92.0);n=57 | 80.0(66.0-100.0);n=817 | 0.6285 |
| Mean ventricular rate | 81.0(68.0-97.0);n=874 | 83.0(76.0-91.0);n=57 | 80.0(68.0-97.0);n=817 | 0.3177 |
| Mean PT interval | 161.0(144.5-177.5);n=874 | 160.0(148.0-178.0);n=57 | 161.0(144.0-177.0);n=817 | 0.649 |
| Mean PR segment | 63.0(48.0-78.0);n=874 | 64.0(48.0-78.0);n=57 | 63.0(49.0-78.0);n=817 | 0.8753 |
| Mean QRS duration | 85.0(78.0-95.0);n=874 | 86.0(81.0-100.0);n=57 | 85.0(78.0-94.0);n=817 | 0.0889 |
| Mean QT interval | 377.0(345.0-408.5);n=874 | 380.0(348.0-415.0);n=57 | 377.0(344.0-408.0);n=817 | 0.5507 |
| Mean QTc interval | 433.0(413.0-458.0);n=874 | 443.0(424.0-464.0);n=57 | 433.0(412.0-458.0);n=817 | 0.0202* |
| QT interval dispersion | 64.0(0.0-104.0);n=874 | 60.0(0.0-104.0);n=57 | 64.0(0.0-104.0);n=817 | 0.686 |

COPD: chronic obstructive pulmonary disease; ACEI: angiotensinogen converting enzyme inhibitor; ARB: angiotensin receptor blocker; APTT: activated partial thromboplastin time.
\* for $p \leq 0.05$ , \*\* for $p \leq 0.01$ , \*\*\* for $p \leq 0.001$ .

**Table 2.**
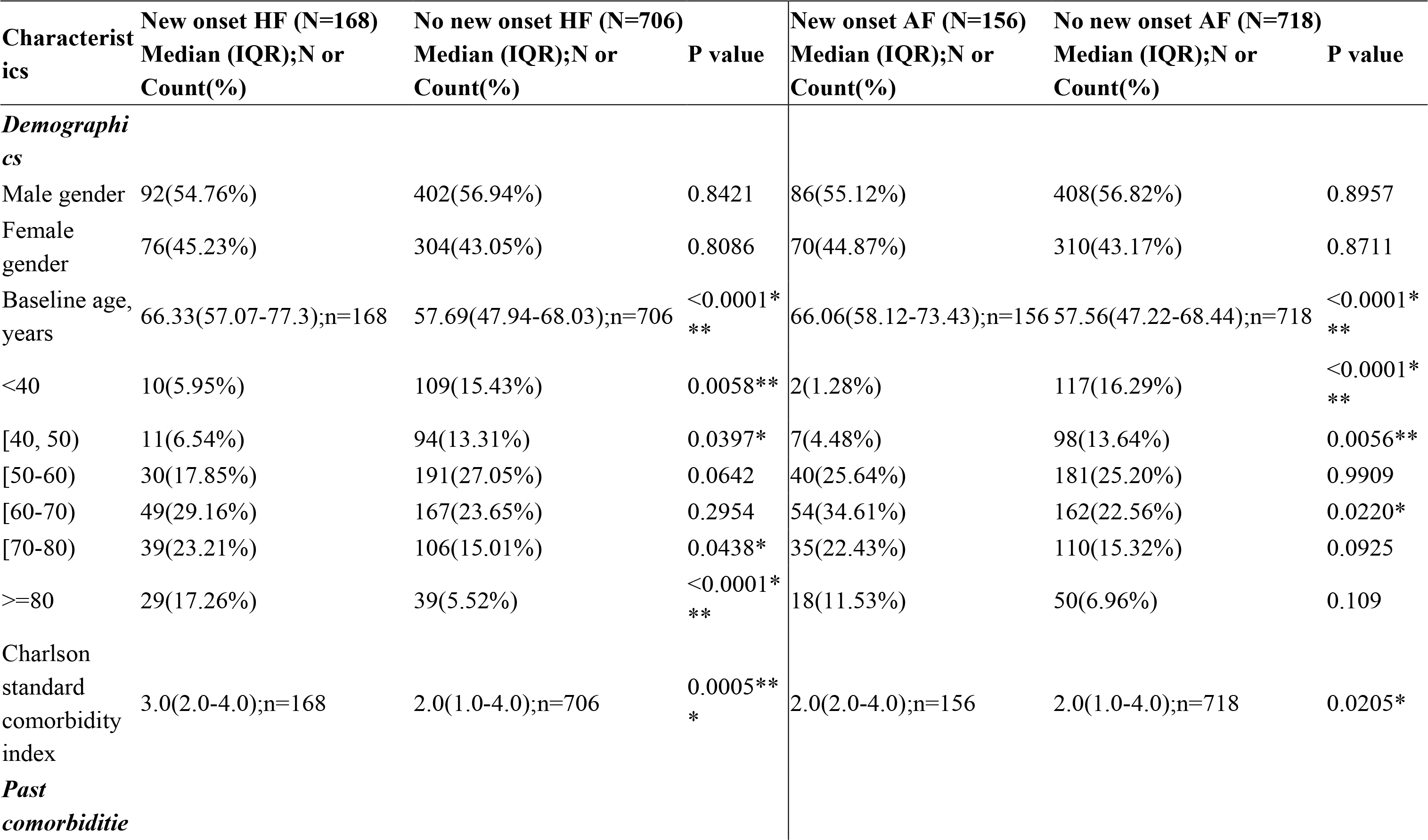
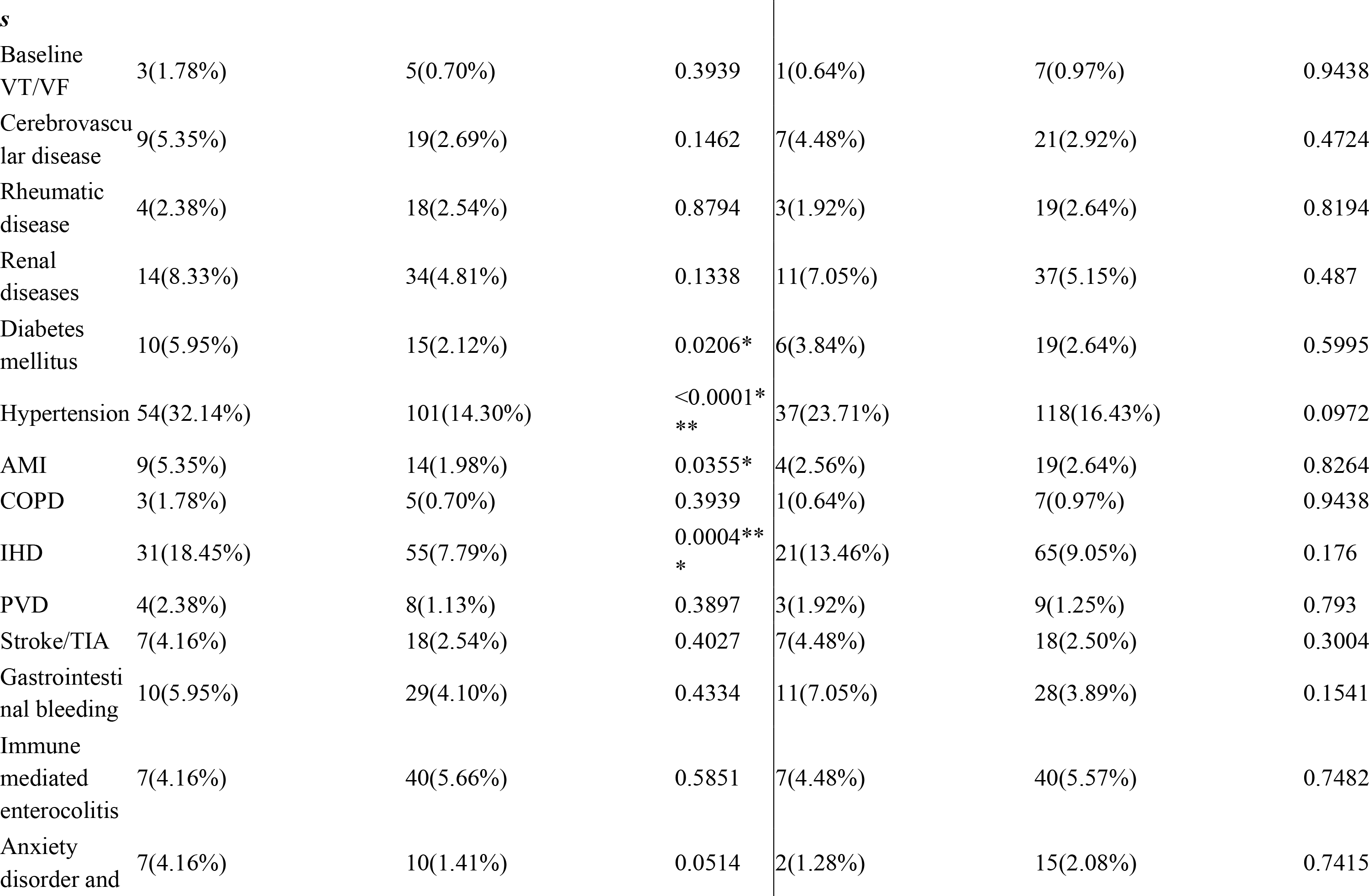
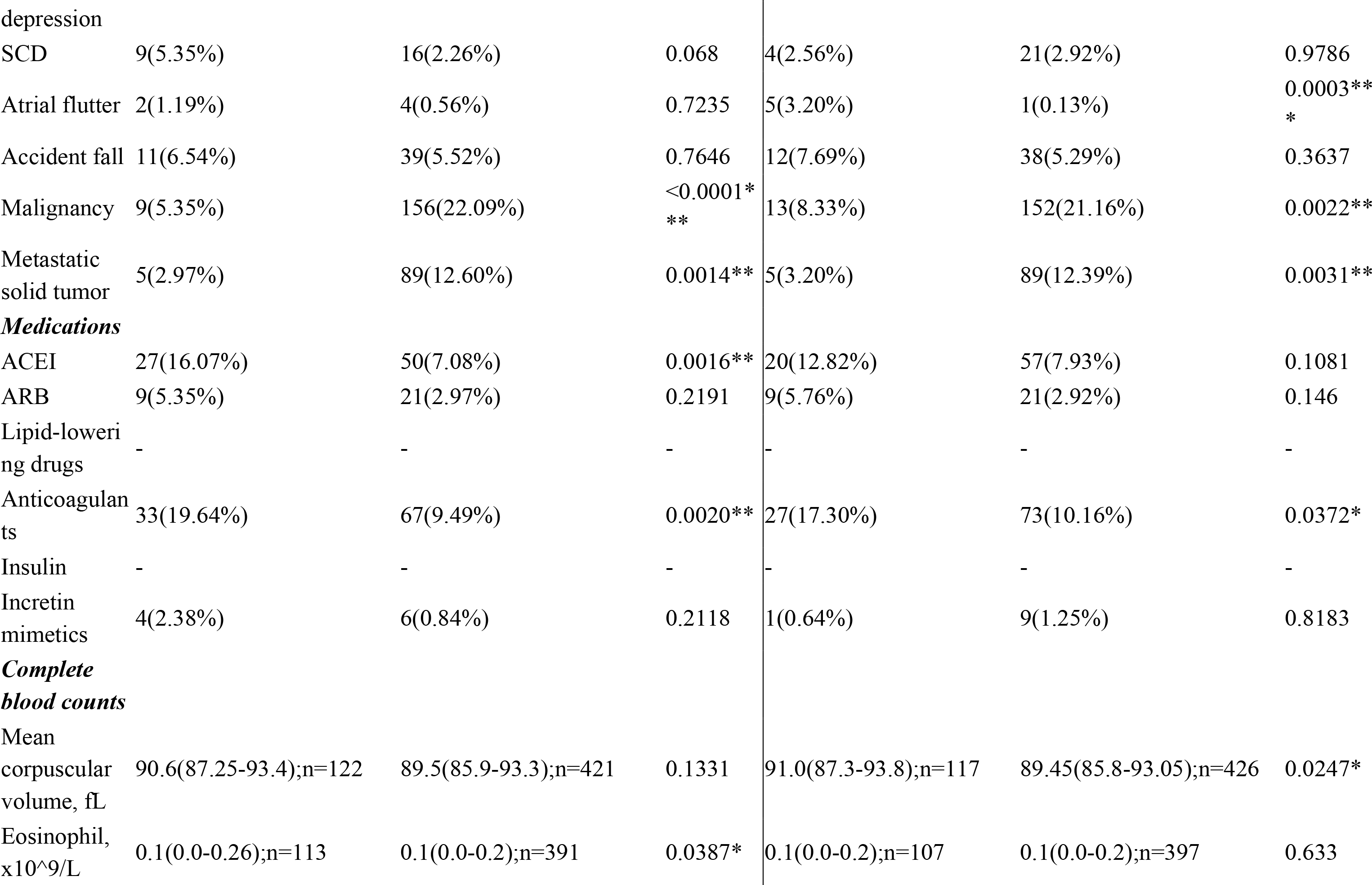
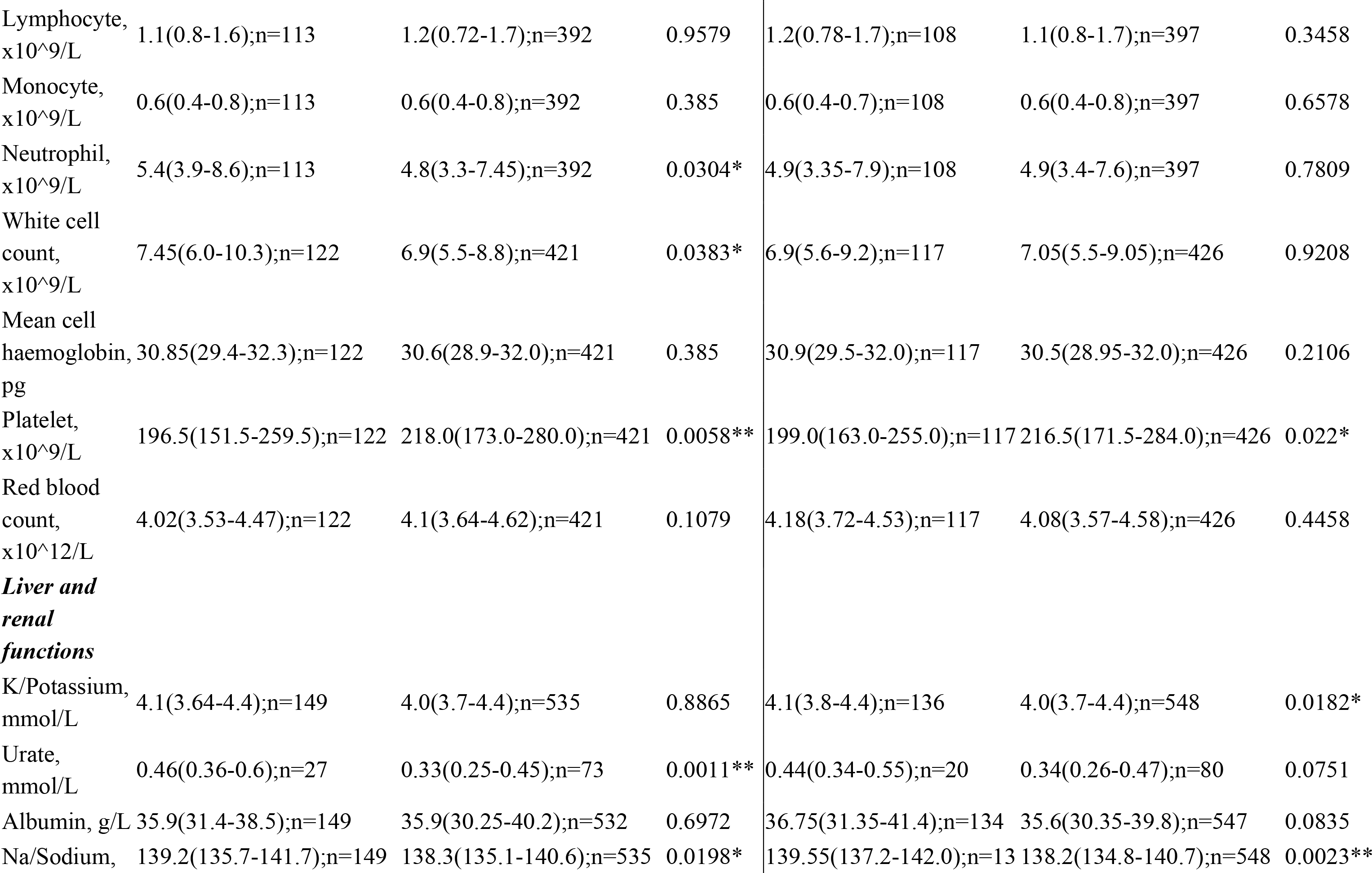
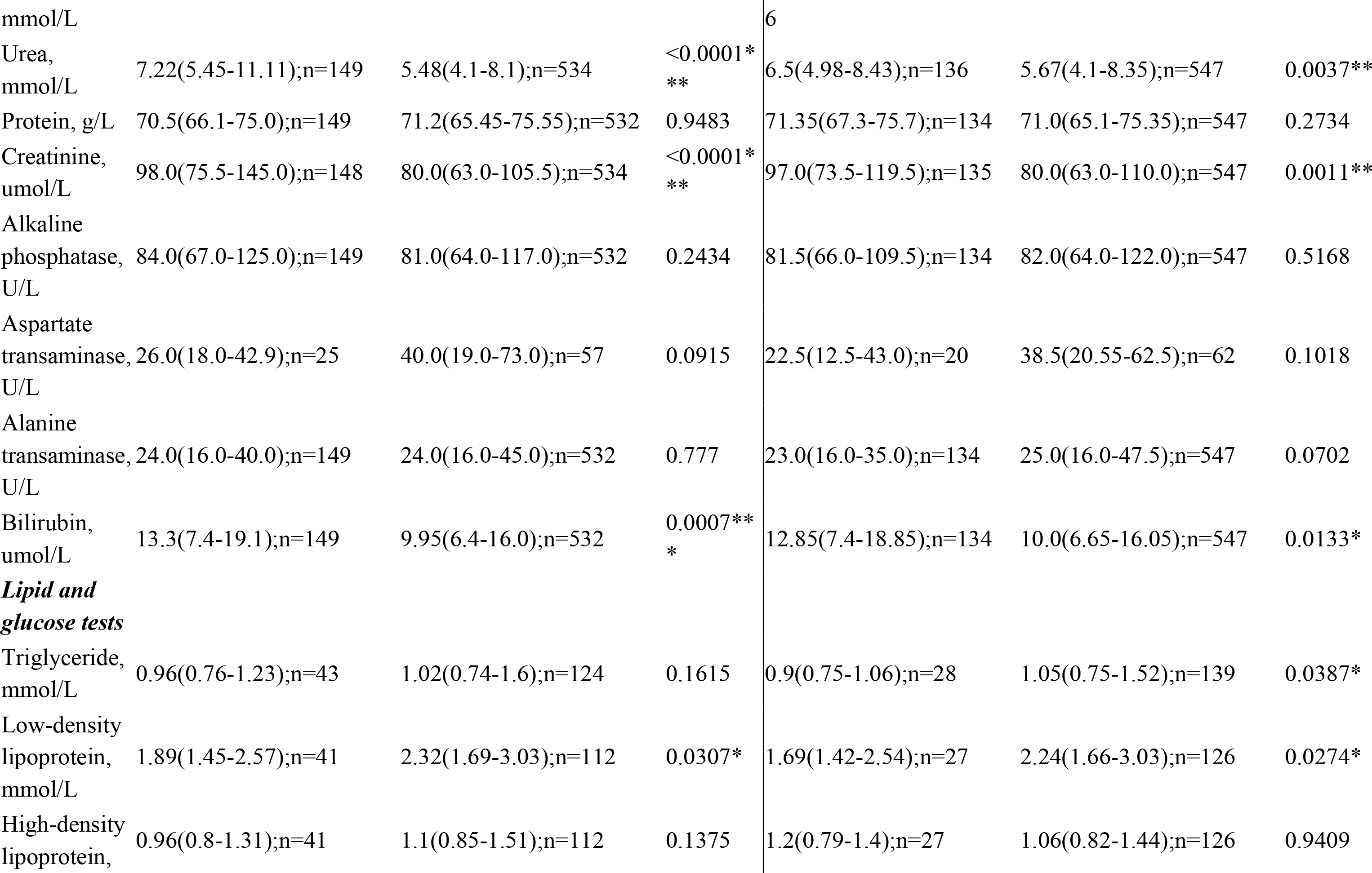
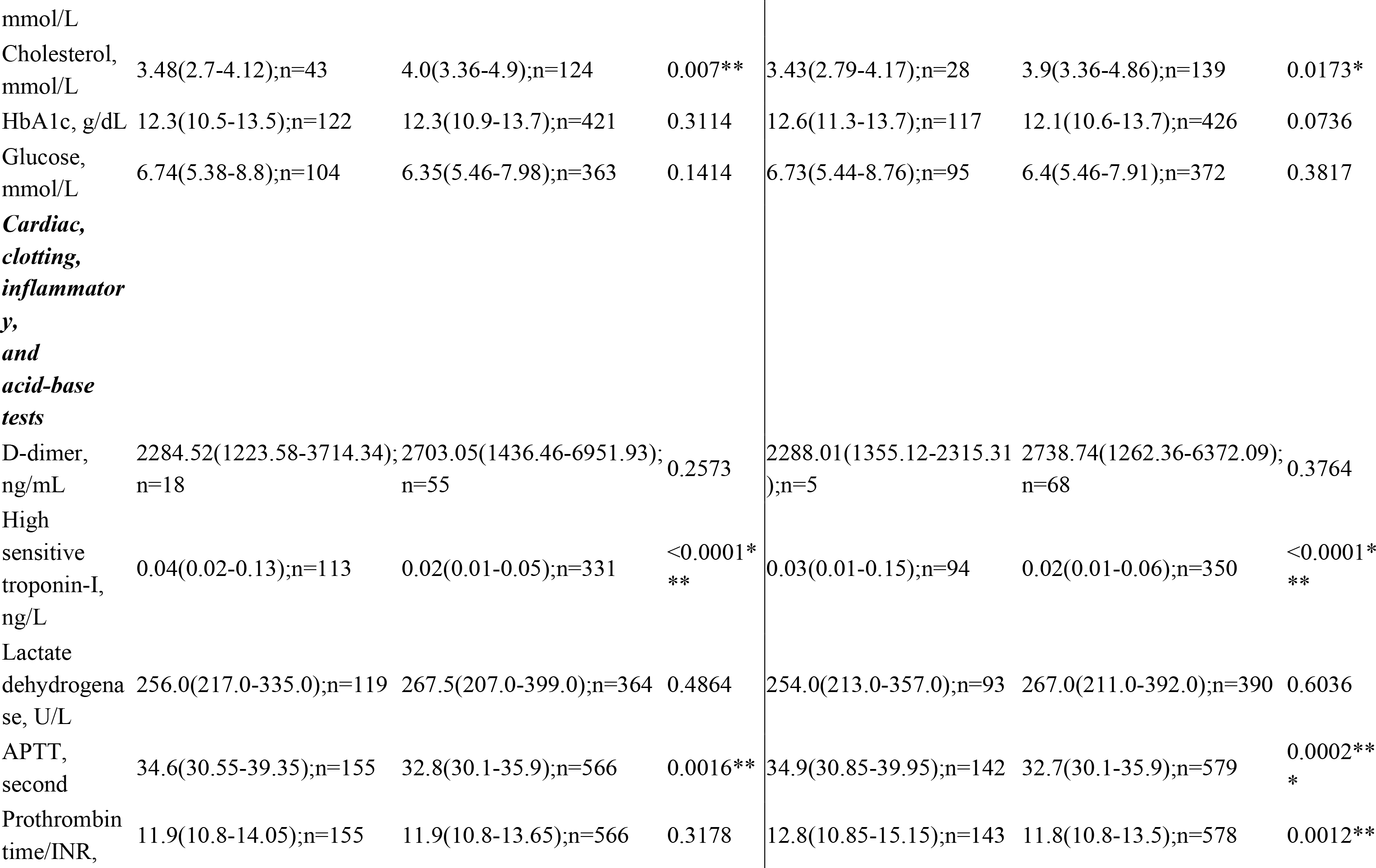
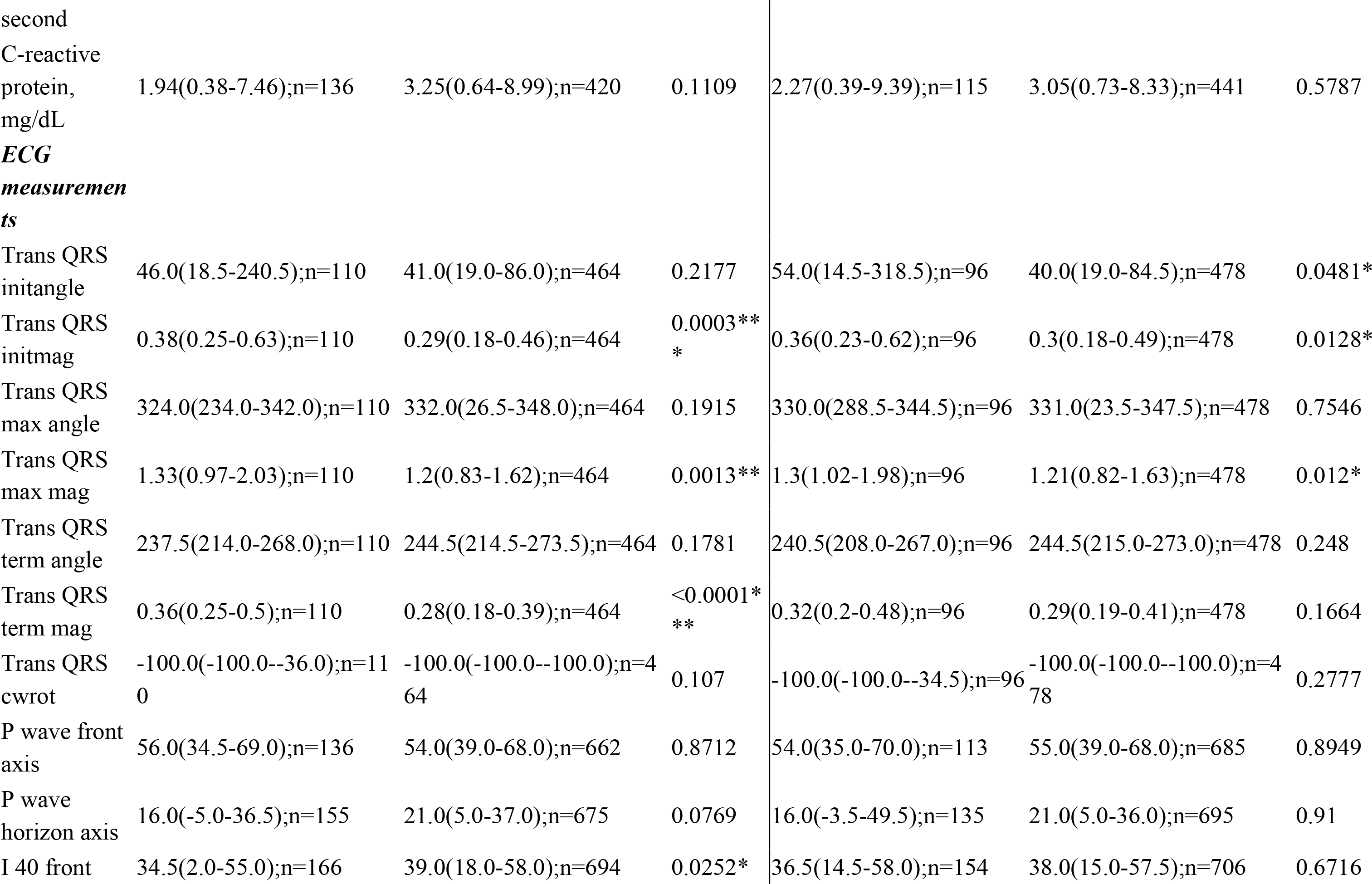
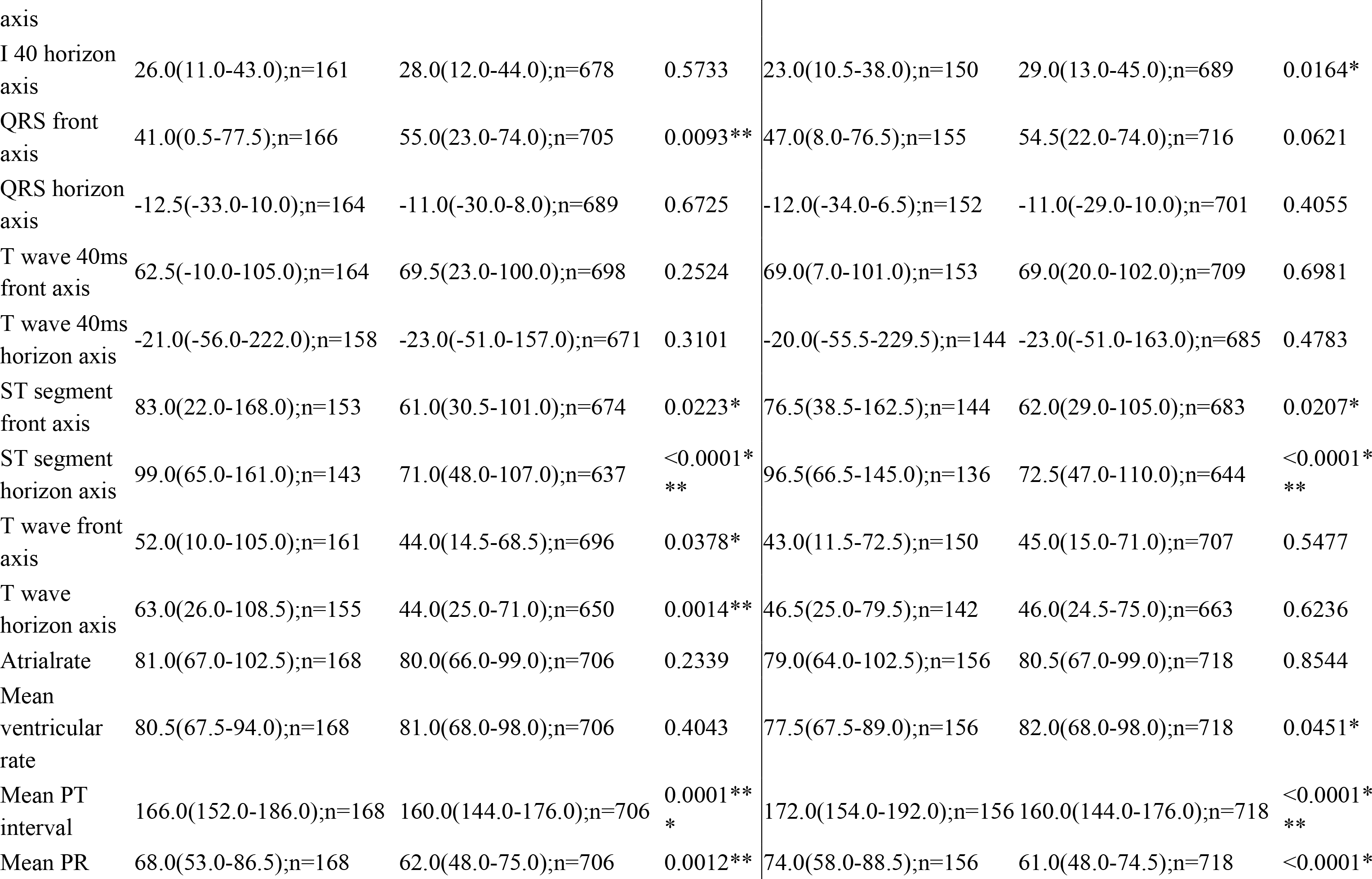
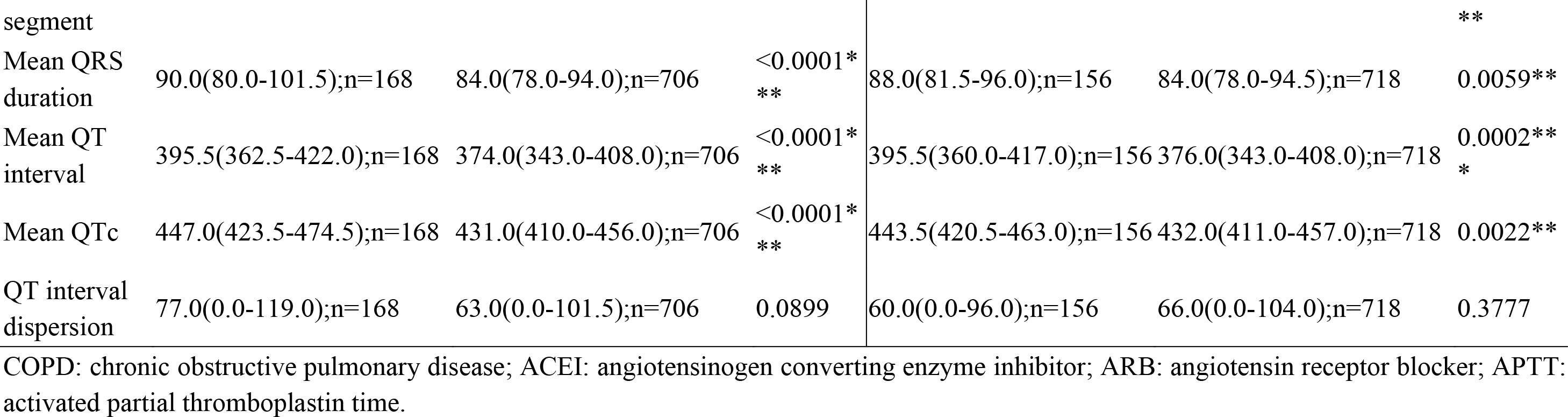
Baseline and clinical characteristics of patients with new onset HF and new onset AF. * for p≤ 0.05, ** for p ≤ 0.01, *** for p ≤ 0.001.

**Table 3.** Univariate Cox analysis of significant risk factors to predict spontaneous VT/VF, new onset HF and new onset AF. * for p≤ 0.05, ** for p ≤ 0.01, *** for p ≤ 0.001

| Characteristics | Spontaneous VT/VF<br>HR [95% CI];P value | Cutoff | New onset HF<br>HR [95% CI];P value | Cutoff | New onset AF<br>HR [95% CI];P value | Cutoff |
| --- | --- | --- | --- | --- | --- | --- |
| ***Demographics*** |  |  |  |  |  |  |
| Male gender | 0.84[0.50-1.42];0.5177 | - | 0.91[0.67-1.25];0.5643 | - | 0.95[0.68-1.31];0.7399 | - |
| Female gender | 1.19[0.70-2.01];0.5177 | - | 1.10[0.80-1.50];0.5643 | - | 1.06[0.76-1.46];0.7399 | - |
| Baseline age, years | 1.00[0.98-1.01];0.6879 | 35.13 | 1.04[1.03-1.05];<0.0001*** | 77.11 | 1.04[1.03-1.06];<0.0001*** | 49.46 |
| <40 | 1.55[0.82-2.93];0.1808 | - | 0.35[0.18-0.66];0.0013** | - | 0.07[0.02-0.27];0.0001*** | - |
| [40, 50) | 0.64[0.26-1.60];0.3409 | - | 0.42[0.22-0.80];0.0085** | - | 0.29[0.14-0.63];0.0016** | - |
| [50-60) | 0.86[0.46-1.61];0.6421 | - | 0.58[0.38-0.87];0.0094** | - | 0.98[0.68-1.42];0.9185 | - |
| [60-70) | 1.09[0.59-1.99];0.7835 | - | 1.45[1.03-2.03];0.0328* | - | 1.74[1.24-2.45];0.0015** | - |
| [70-80) | 1.19[0.60-2.36];0.6219 | - | 1.62[1.11-2.35];0.0121* | - | 1.87[1.28-2.74];0.0013** | - |
| $\geq 80$ | 0.50[0.12-2.06];0.3382 | - | 3.40[2.27-5.10];<0.0001*** | - | 1.93[1.18-3.16];0.0093** | - |
| ***Past comorbidities*** |  |  |  |  |  |  |
| Baseline VT/VF | 13.06[4.70-36.28];<0.0001*** | - | 2.12[0.68-6.65];0.1967 | - | 0.66[0.09-4.70];0.6754 | - |
| Cerebrovascular disease | 1.12[0.27-4.58];0.8790 | - | 1.96[1.00-3.84];0.0504 | - | 1.59[0.74-3.39];0.2346 | - |
| Rheumatic disease | 2.21[0.80-6.14];0.1269 | - | 0.84[0.31-2.26];0.7284 | - | 0.56[0.18-1.75];0.3153 | - |
| Renal diseases | 0.88[0.28-2.83];0.8364 | - | 1.55[0.90-2.69];0.1158 | - | 1.17[0.63-2.17];0.6117 | - |
| Diabetes mellitus | 1.99[0.62-6.37];0.2471 | - | 2.55[1.34-4.83];0.0043** | - | 1.50[0.66-3.40];0.3321 | - |
| Hypertension | 1.21[0.62-2.34];0.5752 | - | 2.73[1.96-3.80];<0.0001*** | - | 1.61[1.10-2.35];0.0149* | - |
| Malignant dysrhythmia | 9.05[3.60-22.70];<0.0001*** | - | 1.65[0.61-4.44];0.3254 | - | 1.36[0.43-4.27];0.5977 | - |
| AMI | - | - | 2.51[1.18-5.36];0.0174* | - | 1.19[0.44-3.23];0.7280 | - |
| COPD | 2.10[0.29-15.19];0.4622 | - | 2.27[0.72-7.12];0.1600 | - | 0.72[0.10-5.13];0.7410 | - |
| IHD | 1.04[0.45-2.42];0.9293 | - | 2.04[1.35-3.08];0.0008*** | - | 1.49[0.94-2.36];0.0935 | - |
| PVD | 1.20[0.17-8.69];0.8557 | - | 1.67[0.62-4.50];0.3135 | - | 1.41[0.45-4.43];0.5563 | - |
| Stroke/TIA | 1.22[0.30-5.02];0.7803 | - | 1.64[0.77-3.51];0.1988 | - | 1.78[0.83-3.81];0.1352 | - |
| Gastrointestinal bleeding | 0.90[0.22-3.68];0.8804 | - | 1.58[0.83-2.99];0.1628 | - | 1.59[0.81-3.11];0.1801 | - |
| Immune mediated enterocolitis | 0.62[0.15-2.54];0.5065 | - | 0.74[0.35-1.58];0.4382 | - | 0.70[0.31-1.58];0.3893 | - |
| Anxiety disorder and depression | 0.94[0.13-6.77];0.9478 | - | 2.73[1.28-5.83];0.0095** | - | 0.74[0.18-3.01];0.6789 | - |
| SCD | 0.82[0.11-5.92];0.8431 | - | 2.26[1.06-4.82];0.0355* | - | 1.10[0.41-2.98];0.8475 | - |
| Atrial flutter | - | - | 1.07[0.15-7.62];0.9485 | - | 5.61[2.07-15.20];0.0007*** | - |
| Accident fall | 1.03[0.32-3.30];0.9617 | - | 1.25[0.67-2.30];0.4828 | - | 1.71[0.95-3.08];0.0761 | - |
| Malignancy | 1.04[0.49-2.21];0.9208 | - | 0.32[0.16-0.63];0.0010** | - | 0.50[0.27-0.90];0.0202* | - |
| Metastatic solid tumor | 0.63[0.15-2.62];0.5274 | - | 0.46[0.19-1.12];0.0873 | - | 0.52[0.21-1.28];0.1530 | - |
| **Medications** |  |  |  |  |  |  |
| ACEI | 1.27[0.58-2.81];0.5525 | - | 1.98[1.31-2.99];0.0013** | - | 1.30[0.80-2.11];0.2826 | - |
| ARB | 2.13[0.85-5.35];0.1074 | - | 1.33[0.68-2.60];0.4116 | - | 1.45[0.74-2.85];0.2802 | - |
| Lipid-lowering drugs | - | - | - | - | - | - |
| Anticoagulants | 1.57[0.81-3.05];0.1776 | - | 1.80[1.23-2.65];0.0026** | - | 1.38[0.90-2.11];0.1355 | - |
| Insulin | - | - | - | - | - | - |
| Incretin mimetics | 1.31[0.18-9.50];0.7869 | - | 2.26[0.84-6.11];0.1069 | - | 0.47[0.07-3.33];0.4469 | - |
| **Complete blood counts** |  |  |  |  |  |  |
| Mean corpuscular volume, fL | 0.98[0.94-1.01];0.2003 | 90 | 1.02[0.99-1.04];0.1297 | 95 | 1.02[0.99-1.04];0.1925 | 90.7 |
| Eosinophil, x10<sup>9</sup>/L | 1.52[0.81-2.86];0.1902 | 0.17 | 1.38[0.97-1.97];0.0773 | 0.19 | 0.97[0.53-1.75];0.9094 | 0.09 |
| Lymphocyte, x10<sup>9</sup>/L | 0.82[0.50-1.34];0.4210 | 1.1 | 0.90[0.70-1.15];0.3919 | 1.15 | 0.99[0.78-1.25];0.9186 | 0.6 |
| Monocyte, x10<sup>9</sup>/L | 1.37[0.94-1.99];0.1023 | 0.65 | 1.40[1.13-1.75];0.0025** | 0.49 | 1.03[0.72-1.49];0.8588 | 0.47 |
| Neutrophil, x10<sup>9</sup>/L | 0.98[0.89-1.08];0.7162 | 4.9 | 1.06[1.01-1.11];0.0084** | 4.7 | 1.04[0.99-1.09];0.1068 | 6.3 |
| White cell count, x10<sup>9</sup>/L | 1.07[0.99-1.16];0.0963 | 6.4 | 1.07[1.03-1.12];0.0017** | 8.3 | 1.01[0.96-1.07];0.7197 | 8.2 |
| Mean cell haemoglobin, pg | 0.94[0.86-1.03];0.1745 | 30.6 | 1.02[0.96-1.08];0.4921 | 33.4 | 1.02[0.96-1.08];0.5375 | 30.4 |
| Platelet, x10<sup>9</sup>/L | 1.002[0.998-1.005];0.3552 | 155 | 0.997[0.995-0.999];0.0135* | 215 | 0.998[0.996-1.000];0.0943 | 275 |
| Red blood count, x10<sup>12</sup>/L | 0.83[0.55-1.26];0.3890 | 3.4 | 0.76[0.61-0.95];0.0180* | 3.97 | 1.04[0.82-1.32];0.7469 | 3.62 |
| ***Liver and renal functions*** |  |  |  |  |  |  |
| K/Potassium, mmol/L | 0.90[0.57-1.43];0.6644 | 3.5 | 0.99[0.76-1.30];0.9640 | 3.64 | 1.27[0.98-1.64];0.0716 | 3.8 |
| Urate, mmol/L | 1.92[0.07-50.43];0.6952 | 0.57 | 34.00[3.86-299.69];0.0015** | 0.43 | 3.17[0.25-40.88];0.3769 | 0.41 |
| Albumin, g/L | 0.98[0.94-1.01];0.2225 | 39.8 | 0.98[0.96-1.00];0.1147 | 39.7 | 1.00[0.97-1.02];0.8670 | 25.3 |
| Na/Sodium, mmol/L | 0.99[0.93-1.05];0.7526 | 141.2 | 1.02[0.98-1.06];0.3282 | 141.1 | 1.05[1.00-1.09];0.0366* | 141.8 |
| Urea, mmol/L | 1.01[0.97-1.04];0.7378 | 11.89 | 1.02[1.01-1.03];0.0053** | 6.6 | 1.01[0.99-1.03];0.4214 | 4.11 |
| Protein, g/L | 0.99[0.96-1.02];0.5079 | 77.2 | 0.99[0.97-1.01];0.5958 | 77.6 | 1.00[0.98-1.02];0.9728 | 66.4 |
| Creatinine, umol/L | 1.000[0.999-1.001];0.6363 | 60 | 1.000[1.000-1.001];0.5454 | 122 | 1.000[0.999-1.001];0.9813 | 85 |
| Alkaline phosphatase, U/L | 1.00[1.00-1.01];0.3039 | 67 | 1.001[0.999-1.004];0.3772 | 65 | 1.000[0.997-1.003];0.8691 | 63 |
| Aspartate transaminase, U/L | 1.001[0.999-1.002];0.2601 | 21 | 0.99[0.97-1.00];0.1255 | 49 | 1.000[0.998-1.001];0.6384 | 23 |
| Alanine transaminase, U/L | 1.000[0.999-1.001];0.7930 | 15.9 | 1.001[1.000-1.001];0.0027** | 91 | 1.00[0.99-1.00];0.2503 | 49 |
| Bilirubin, umol/L | 0.99[0.97-1.02];0.6014 | 7.8 | 1.01[1.00-1.01];0.0002*** | 12.3 | 1.01[1.01-1.02];<0.0001*** | 12 |
| ***Lipid and glucose tests*** |  |  |  |  |  |  |
| Triglyceride, mmol/L | 1.76[0.84-3.68];0.1323 | 1.21 | 0.61[0.34-1.11];0.1053 | 1.69 | 0.37[0.15-0.90];0.0281* | 1.23 |
| Low-density lipoprotein, mmol/L | 0.97[0.54-1.74];0.9228 | 1.2109 | 0.66[0.45-0.95];0.0273* | 2.1173 | 0.65[0.40-1.04];0.0724 | 2.7855 |
| High-density lipoprotein, mmol/L | 0.19[0.04-0.94];0.0415* | 1.09 | 0.56[0.28-1.14];0.1109 | 1.56 | 0.96[0.42-2.19];0.9242 | 1.18 |
| Cholesterol, mmol/L | 0.99[0.65-1.52];0.9722 | 4.03 | 0.70[0.53-0.93];0.0126* | 4.7 | 0.68[0.47-0.97];0.0343* | 3.6 |
| HbA1c, g/dL | 0.90[0.78-1.04];0.1583 | 11.4 | 0.92[0.85-1.00];0.0464* | 9.7 | 1.03[0.95-1.13];0.4444 | 10.8 |
| Glucose, mmol/L | 0.96[0.86-1.07];0.4460 | 5.17 | 1.05[1.01-1.09];0.0165* | 6.64 | 1.03[0.98-1.07];0.2317 | 7.79 |
| ***Cardiac, clotting, inflammatory, and acid-base tests*** |  |  |  |  |  |  |
| D-dimer, ng/mL | 1.000[1.000-1.000];0.8538 | 1676.12 | 1.000[1.000-1.000];0.2234 | 6329.5498 | 1.000[1.000-1.000];0.9549 | 2315.31 |
| High sensitive troponin-I, ng/L | 0.99[0.90-1.08];0.7459 | 0.0199 | 1.01[1.00-1.02];0.1592 | 0.0179 | 1.00[0.98-1.02];0.9476 | 0.0114 |
| Lactate dehydrogenase, U/L | 1.001[1.000-1.002];0.0554 | 409 | 1.000[0.999-1.001];0.8341 | 218 | 1.001[1.000-1.001];0.1068 | 223 |
| APTT, second | 1.01[0.99-1.03];0.3782 | 33.9 | 1.01[1.00-1.02];0.0073** | 39.5 | 1.02[1.01-1.03];<0.0001*** | 39.5 |
| Prothrombin time/INR, second | 1.00[0.96-1.05];0.8719 | 10.1 | 1.03[1.01-1.05];0.0030** | 16.1 | 1.04[1.02-1.06];0.0002*** | 12.7 |
| C-reactive protein, mg/dL | 1.01[0.97-1.05];0.5465 | 1.739 | 1.00[0.97-1.02];0.8999 | 4.175 | 1.01[0.99-1.03];0.4155 | 5.975 |
| ***ECG measurements*** |  |  |  |  |  |  |
| Trans QRS initangle | 1.000[0.997-1.003];0.8486 | 21 | 1.002[1.001-1.004];0.0037** | 205 | 1.003[1.001-1.004];0.0002*** | 251 |
| Trans QRS initmag | 0.66[0.18-2.40];0.5259 | 0.11 | 2.77[1.67-4.60];0.0001*** | 0.75 | 2.03[1.09-3.77];0.0257* | 0.582 |
| Trans QRS max angle | 1.000[0.997-1.002];0.7373 | 220 | 1.001[0.999-1.002];0.3282 | 351 | 1.001[1.000-1.003];0.0752 | 7 |
| Trans QRS max mag | 0.70[0.39-1.24];0.2230 | 1.322 | 1.43[1.08-1.90];0.0131* | 2.16 | 1.23[0.91-1.67];0.1847 | 1.97 |
| Trans QRS term angle | 1.00[0.99-1.00];0.0444* | 234 | 1.00[0.99-1.00];0.1175 | 225 | 0.999[0.996-1.002];0.5706 | 257 |
| Trans QRS term mag | 1.74[0.55-5.46];0.3432 | 0.592 | 2.70[1.55-4.68];0.0004*** | 0.49 | 1.58[0.76-3.30];0.2220 | 0.45 |
| Trans QRS cwrot | 1.00[1.00-1.01];0.5411 | -34 | 1.00[1.00-1.01];0.2545 | -50 | 1.00[1.00-1.01];0.4167 | -45 |
| P wave front axis | 1.00[0.99-1.01];0.7611 | 75 | 1.00[1.00-1.01];0.6109 | 15 | 1.00[1.00-1.01];0.2615 | 75 |
| P wave horizon axis | 1.00[0.99-1.00];0.2539 | -18 | 1.00[1.00-1.01];0.2775 | -15 | 1.01[1.00-1.01];0.0002*** | 53 |
| I 40 front axis | 1.00[0.99-1.00];0.3288 | -4 | 0.99[0.99-1.00];0.0242* | 7 | 1.00[0.99-1.00];0.1402 | 26 |
| I 40 horizon axis | 1.01[1.00-1.01];0.0587 | -3 | 1.00[1.00-1.01];0.2961 | 29 | 0.99[0.99-1.00];0.0172* | 29 |
| QRS front axis | 1.00[0.99-1.00];0.5589 | -12 | 0.99[0.99-1.00];0.0017** | 2 | 1.00[0.99-1.00];0.0128* | -2 |
| QRS horizon axis | 1.00[1.00-1.01];0.7183 | -48 | 1.002[0.999-1.004];0.2684 | -50 | 0.999[0.995-1.002];0.4450 | -30 |
| T wave 40ms front axis | 1.001[0.998-1.005];0.4056 | 103 | 0.999[0.996-1.001];0.2277 | -8 | 0.999[0.997-1.002];0.5548 | -41 |
| T wave 40ms horizon axis | 1.002[1.000-1.004];0.1059 | 15 | 1.002[1.000-1.003];0.0131* | 91 | 1.001[1.000-1.002];0.0744 | 226 |
| ST segment front axis | 1.001[0.997-1.004];0.7597 | 105 | 1.002[1.000-1.004];0.0214* | 108 | 1.00[1.00-1.01];0.0011** | 106 |
| ST segment horizon axis | 1.00[1.00-1.01];0.2457 | 149 | 1.01[1.00-1.01];<0.0001*** | 162 | 1.01[1.00-1.01];0.0001*** | 90 |
| T wave front axis | 1.00[1.00-1.01];0.1033 | 22 | 1.00[1.00-1.01];0.0032** | 80 | 1.000[0.997-1.002];0.8900 | 28 |
| T wave horizon axis | 1.000[0.996-1.005];0.8465 | 16 | 1.00[1.00-1.01];0.0208* | 59 | 1.000[0.997-1.003];0.8419 | 70 |
| Atrialrate | 1.00[0.99-1.00];0.4626 | 71 | 1.00[1.00-1.01];<0.0001*** | 116 | 1.01[1.00-1.01];<0.0001*** | 118 |
| Mean ventricular rate | 1.01[1.00-1.02];0.1893 | 77 | 1.00[1.00-1.01];0.4141 | 78 | 1.00[0.99-1.01];0.9446 | 103 |
| Mean PT interval | 1.00[0.99-1.01];0.8872 | 162 | 1.01[1.00-1.01];0.0006*** | 195 | 1.01[1.01-1.02];<0.0001*** | 176 |
| Mean PR segment | 1.00[0.98-1.01];0.4562 | 56 | 1.01[1.00-1.01];0.0046** | 90 | 1.01[1.01-1.02];<0.0001*** | 76 |
| Mean QRS duration | 1.01[1.00-1.03];0.0171* | 93 | 1.02[1.01-1.02];<0.0001*** | 103 | 1.01[1.00-1.01];0.0666 | 77 |
| Mean QT interval | 1.00[0.99-1.01];0.9214 | 338 | 1.01[1.00-1.01];0.0011** | 391 | 1.00[1.00-1.01];0.0051** | 392 |
| Mean QTc | 1.01[1.00-1.02];0.0035** | 413 | 1.01[1.01-1.02];<0.0001*** | 466 | 1.01[1.00-1.01];0.0001*** | 445 |
| QT interval dispersion | 1.001[0.997-1.005];0.7497 | 34 | 1.00[1.00-1.01];0.0004*** | 77 | 1.002[1.000-1.005];0.0868 | 43 |

COPD: chronic obstructive pulmonary disease; ACEI: angiotensinogen converting enzyme inhibitor; ARB: angiotensin receptor blocker; APTT: activated partial thromboplastin time.

**Table 4.**
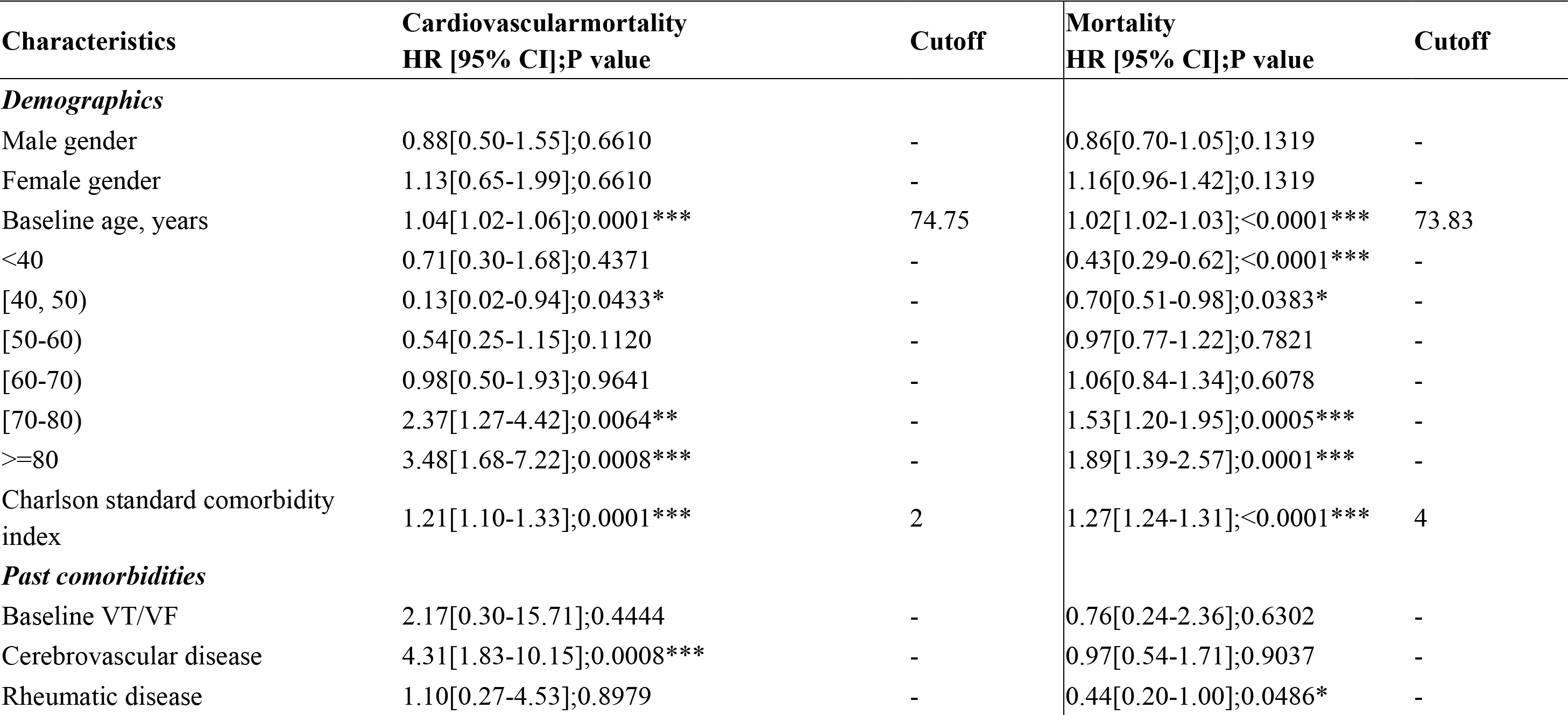
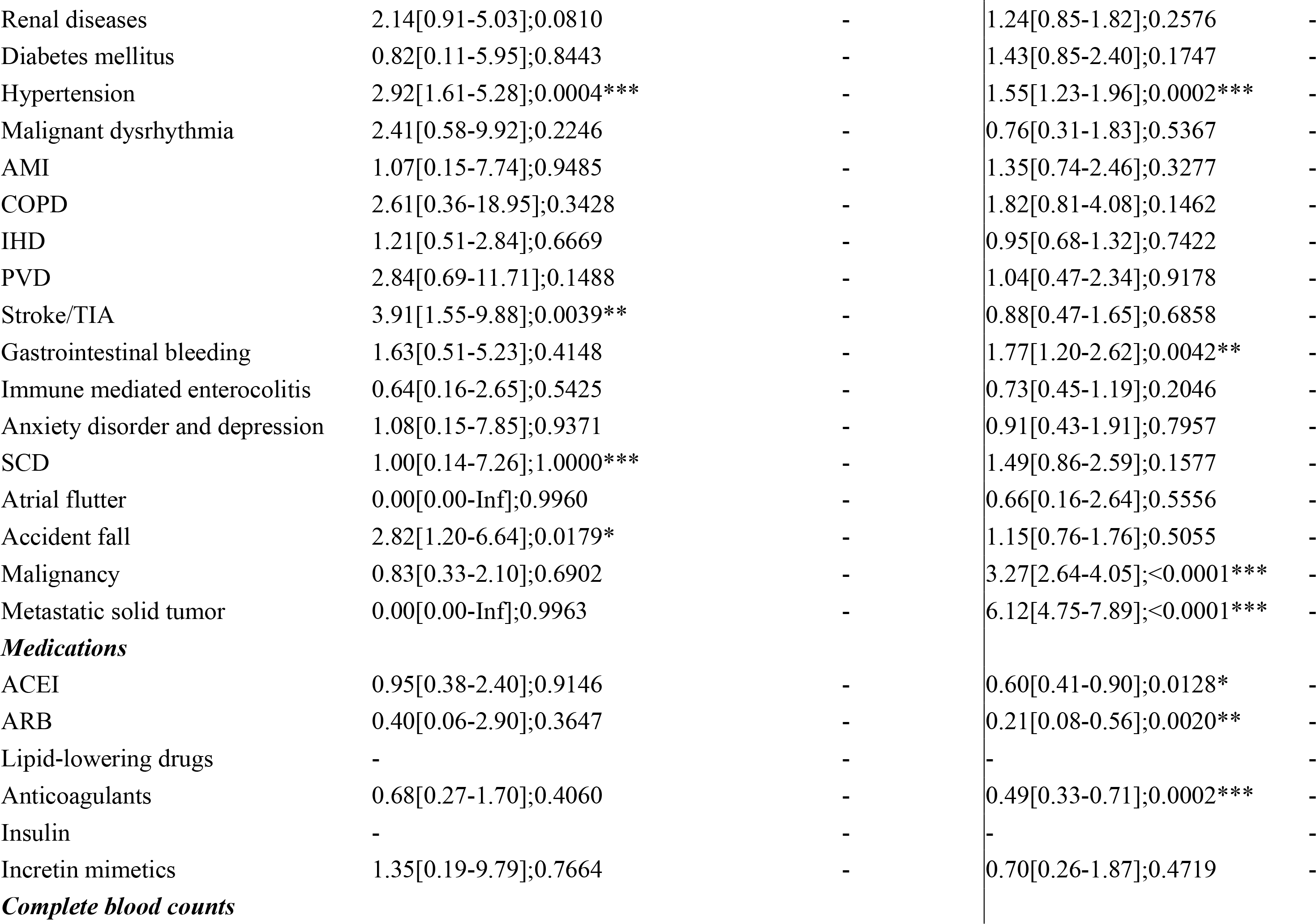
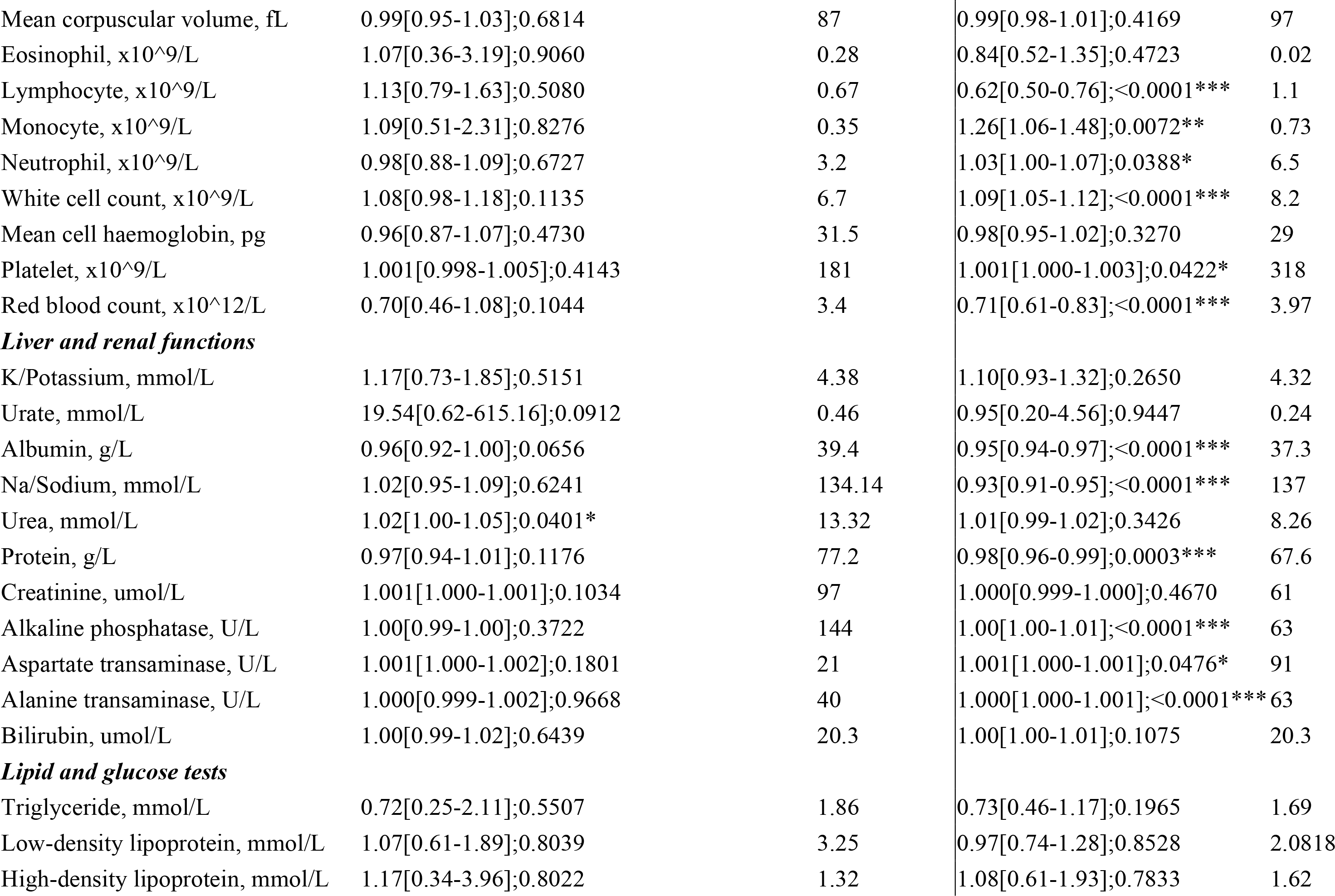
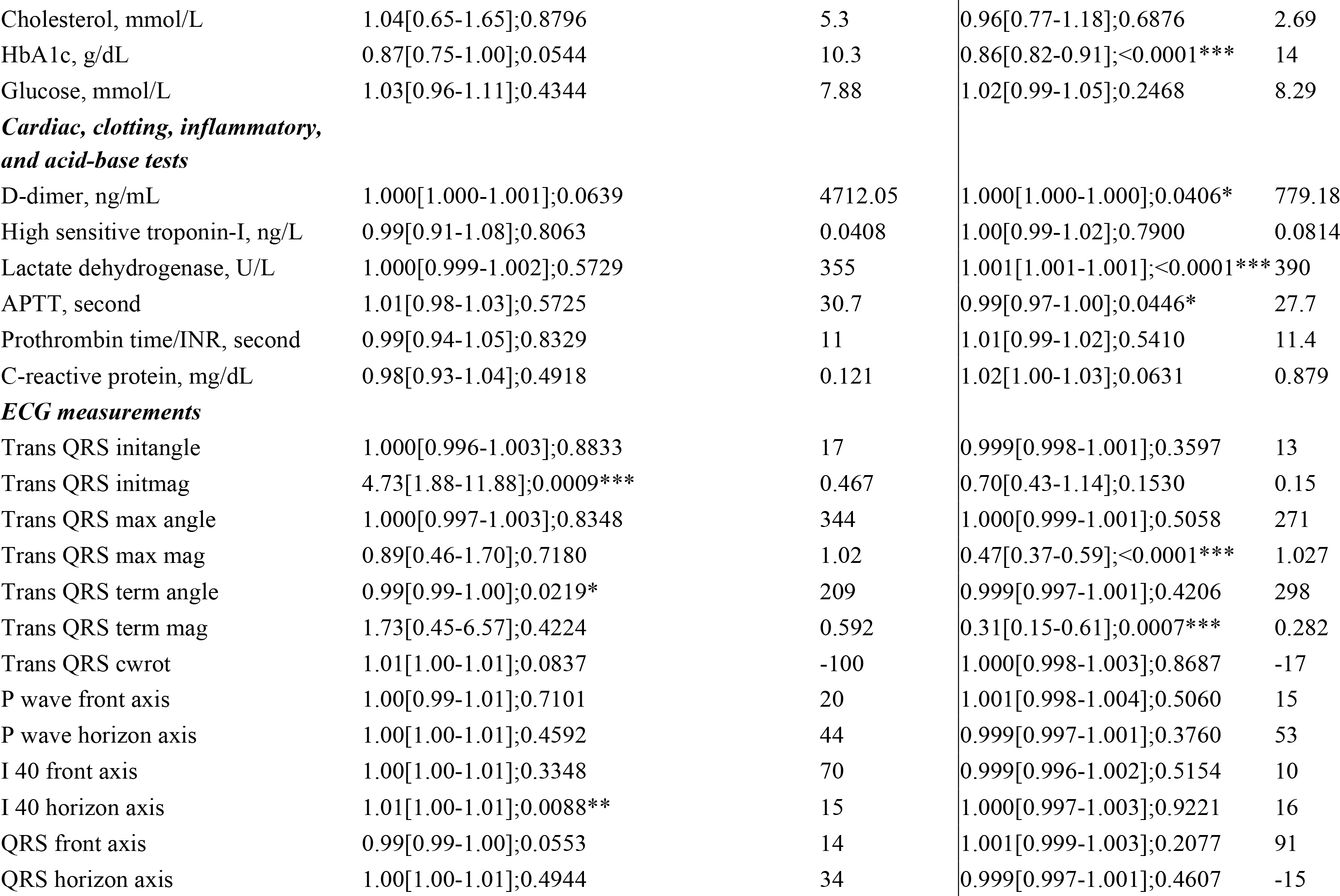
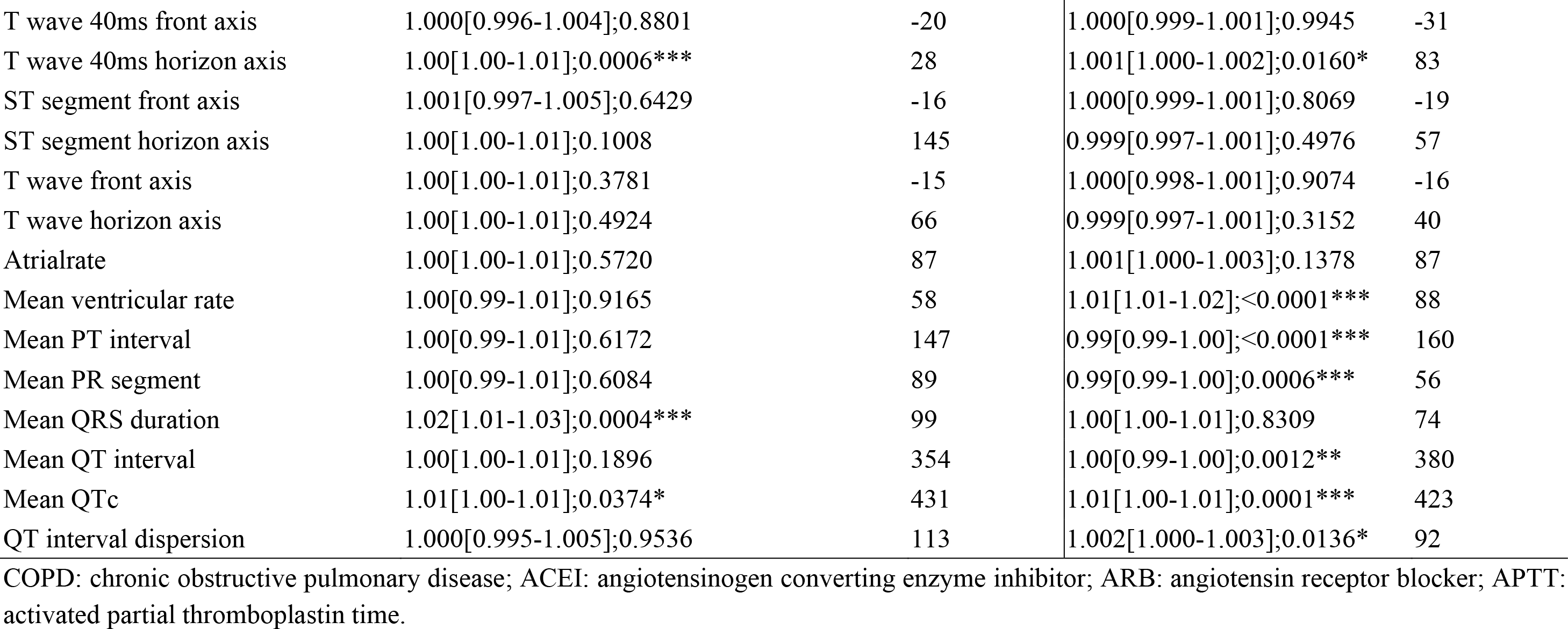
Univariate Cox analysis of significant risk factors to predict cardiovascular mortality and all-cause mortality. * for p≤ 0.05, ** for p ≤ 0.01, *** for p ≤ 0.001

### Predictors of primary and secondary outcomes

A total of 874 patients were included. The cohort was 57% male and had a median age of 59 (IQR: 50-70) years old. During follow-up, 57 patients (6.5%), 156 (17.8%) and 168 (19.2%) suffered from VT/VF, AF and HF, respectively. Cox regression identified baseline VT/VF, terminal angle of the QRS vector in the transverse plane, mean QRS duration and mean QTc intervals as significant predictors of incident VT/VF events, with only the former most maintaining significance in multivariate analysis (HR: 5.03; 95% CI: 1.12 – 22.6; P = 0.035). In contrast, regarding secondary outcomes, baseline age, prior diagnoses of hypertension, initial angle and magnitude of the QRS vector in the transverse plane, P-wave and QRS axis in the frontal plane, ST segment axis in the frontal and horizontal planes, mean PT interval, mean PR segment duration and QTc intervals were all univariate predictors of incident AF, albeit only baseline age (HR: 1.03; 95% CI: 1.02 – 1.05; P < 0.001) and initial angle of the QRS vector in the transverse plane (HR: 1.003; 95% CI: 1.001 – 1.005; P = 0.003) retained significance after multivariate adjustment. As it pertains to new-onset HF, several clinical and electrocardiographic parameters demonstrated an association with HF in univariate analysis, including prior diagnosis of HT (HR: 3.27; 95% CI: 2.08 – 5.13; P < 0.001) or DM, initial QRS angle in transverse plane (HR: 1.002; 95% CI: 1.001 – 1.004; P = 0.008), I 40 in horizontal axis, ST-segment axis in the horizontal plane (HR: 1.004; 95% CI: 1.001 – 1.009; P = 0.008), T-wave frontal axis and atrial rate (HR: 1.01; 95% CI: 1.006 – 1.014; P < 0.001), of which, except for prior diagnosis of DM, I40 in horizontal axis and T-wave frontal axis, all variables showcased significant relationships in multivariate analysis.

A total of 396 (45%) patients died during follow-up, of which 52 (6%) were cardiovascular-related. Univariate predictors for all-cause mortality comprised baseline age, prior diagnosis of hypertension, terminal and maximum magnitude of the QRS vector in the transverse plane, T wave (40ms) horizontal axis, ventricular rate mean QTc interval, mean PT interval, mean PR segment and QT interval dispersion. Subsequent multivariate adjustment revealed only significant associations for only baseline age (HR: 1.02; 95% CI: 1.01-1.03; P < 0.001) and maximum magnitude of the QRS vector in the transverse plane (HR: 0.55; 95% CI: 0.4-0.75; P <0.001).

## Discussion

The major limitation of this study is that it is based on a single centre cohort. We recognize that the derivation populations may differ from other populations based both on hospital conditions and inherent demographic differences. An external validation using independent external cohort allows us to assess the broader clinical utility of our findings. However, the model should be further externally validated using patient data from other regions.

## Data Availability

All data produced in the present study are available upon reasonable request to the authors

---

**Figure 1.**
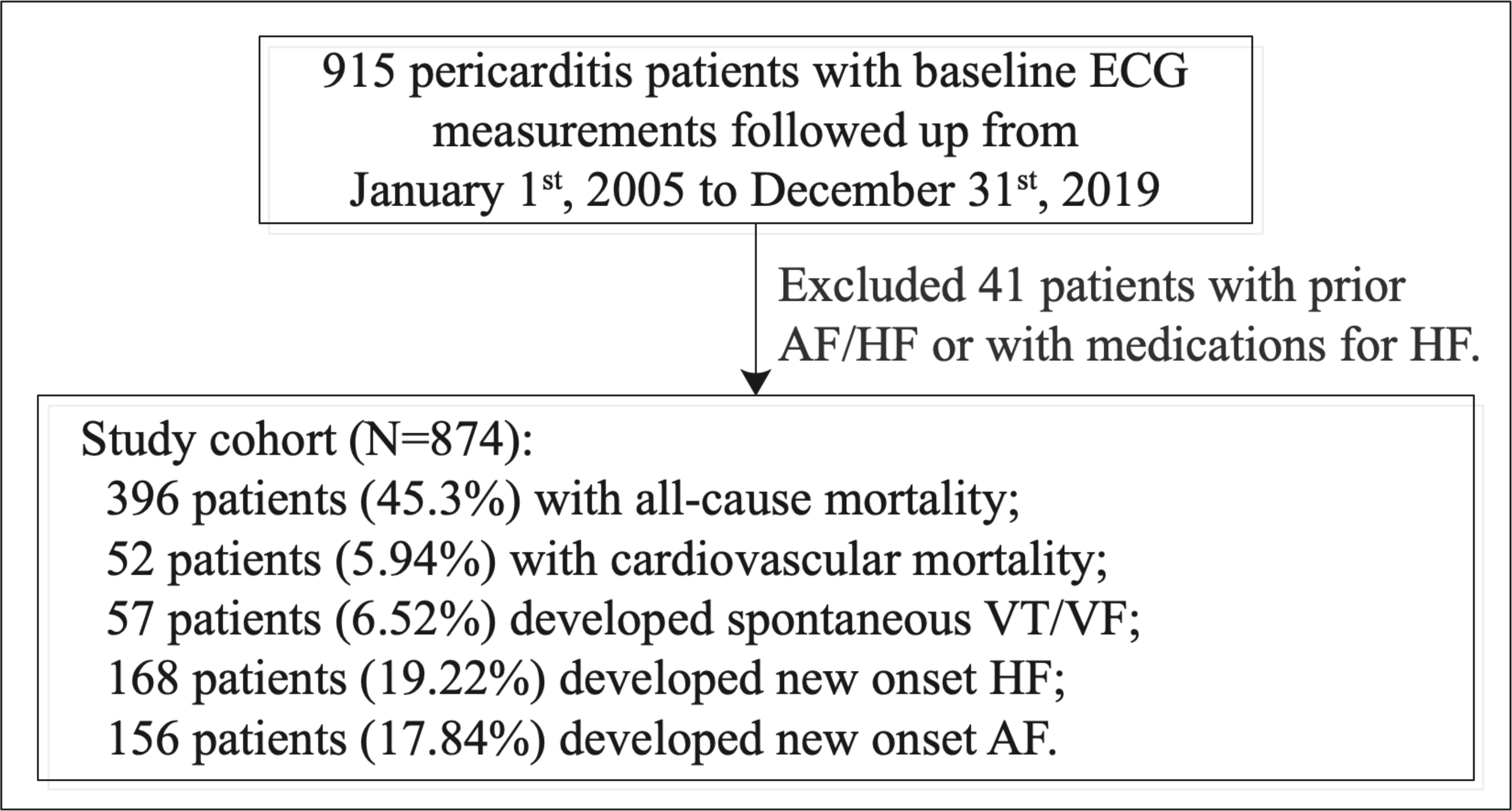
Procedures of data processing.

**Figure 2.**
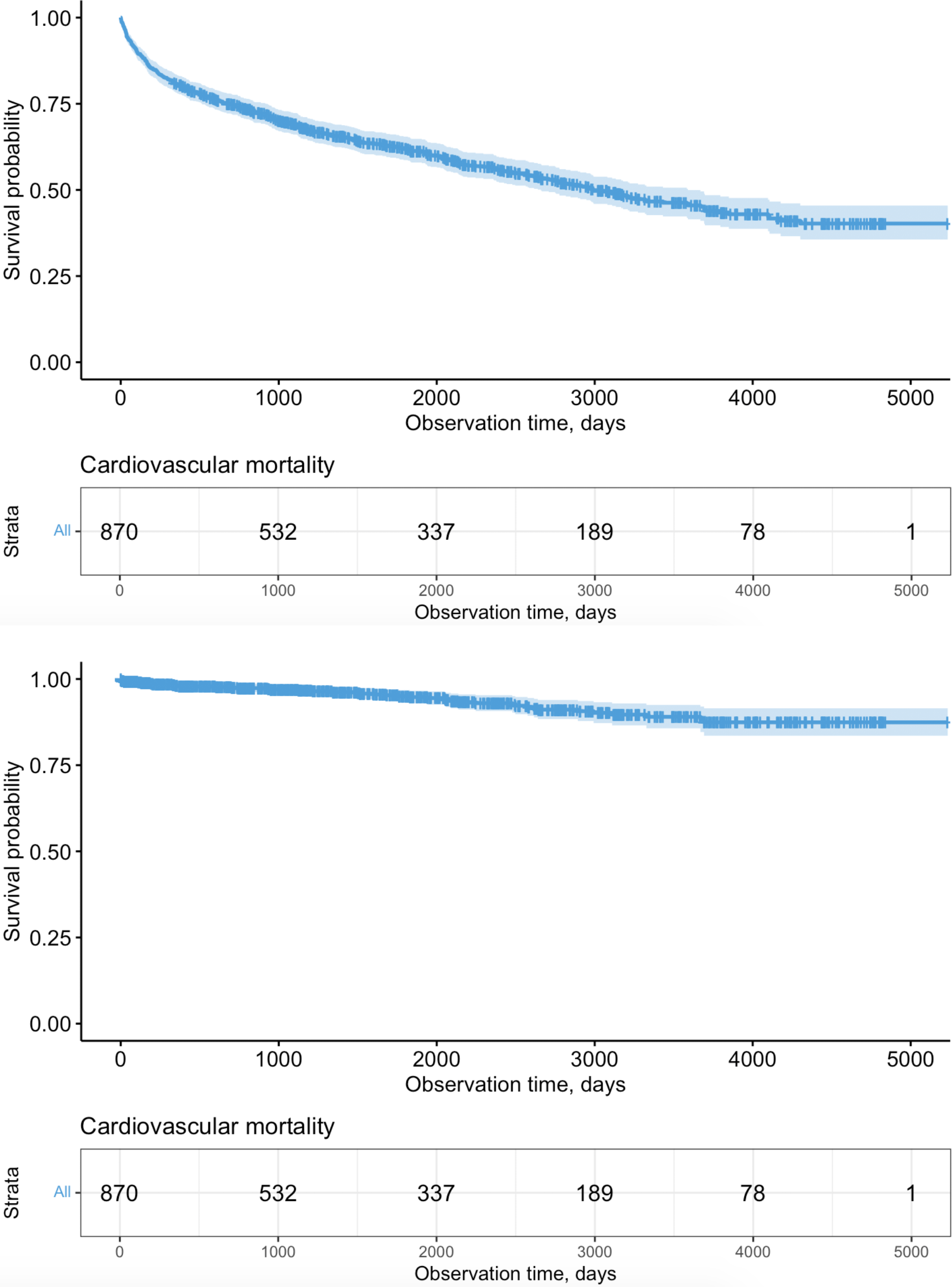
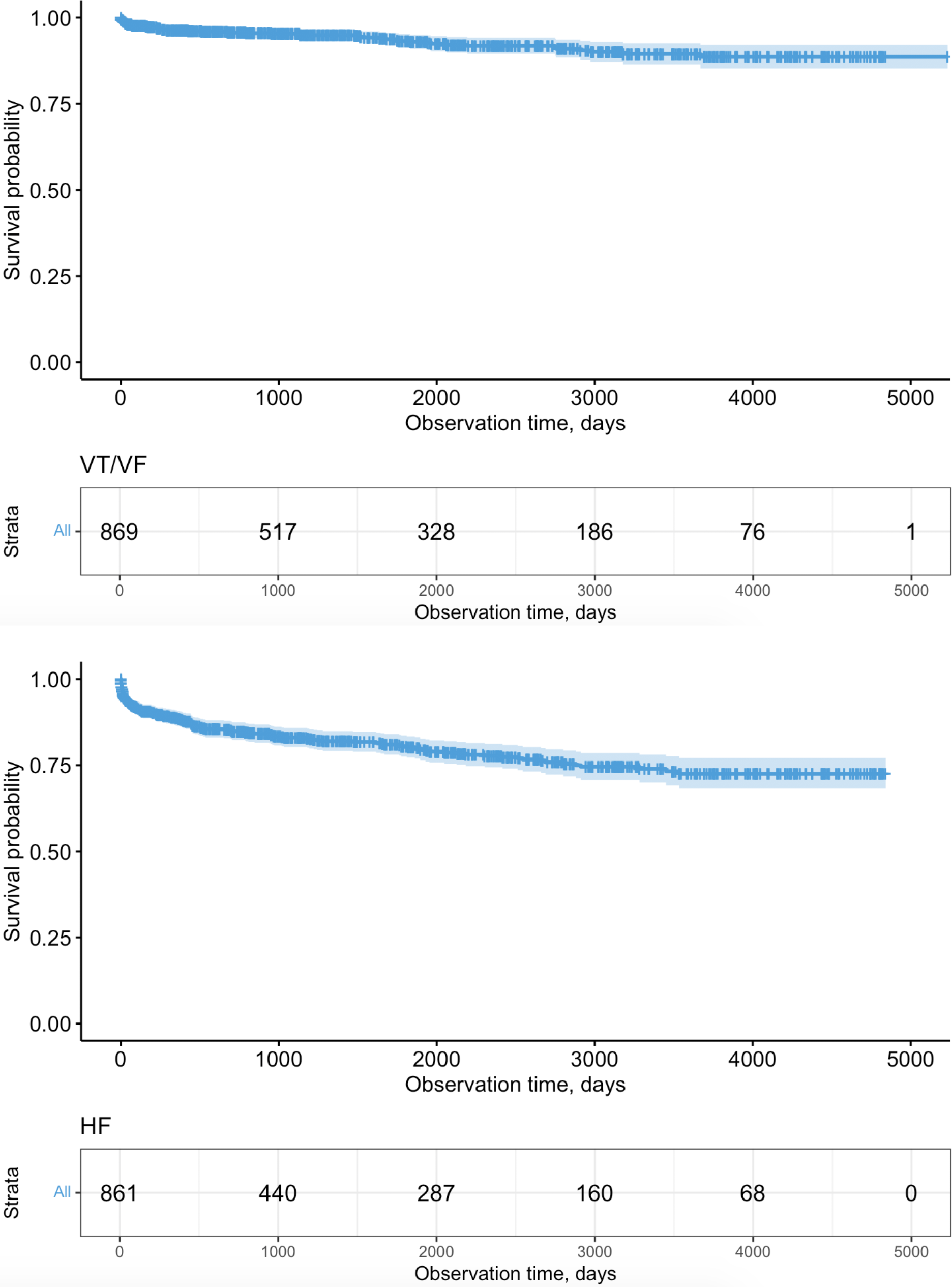
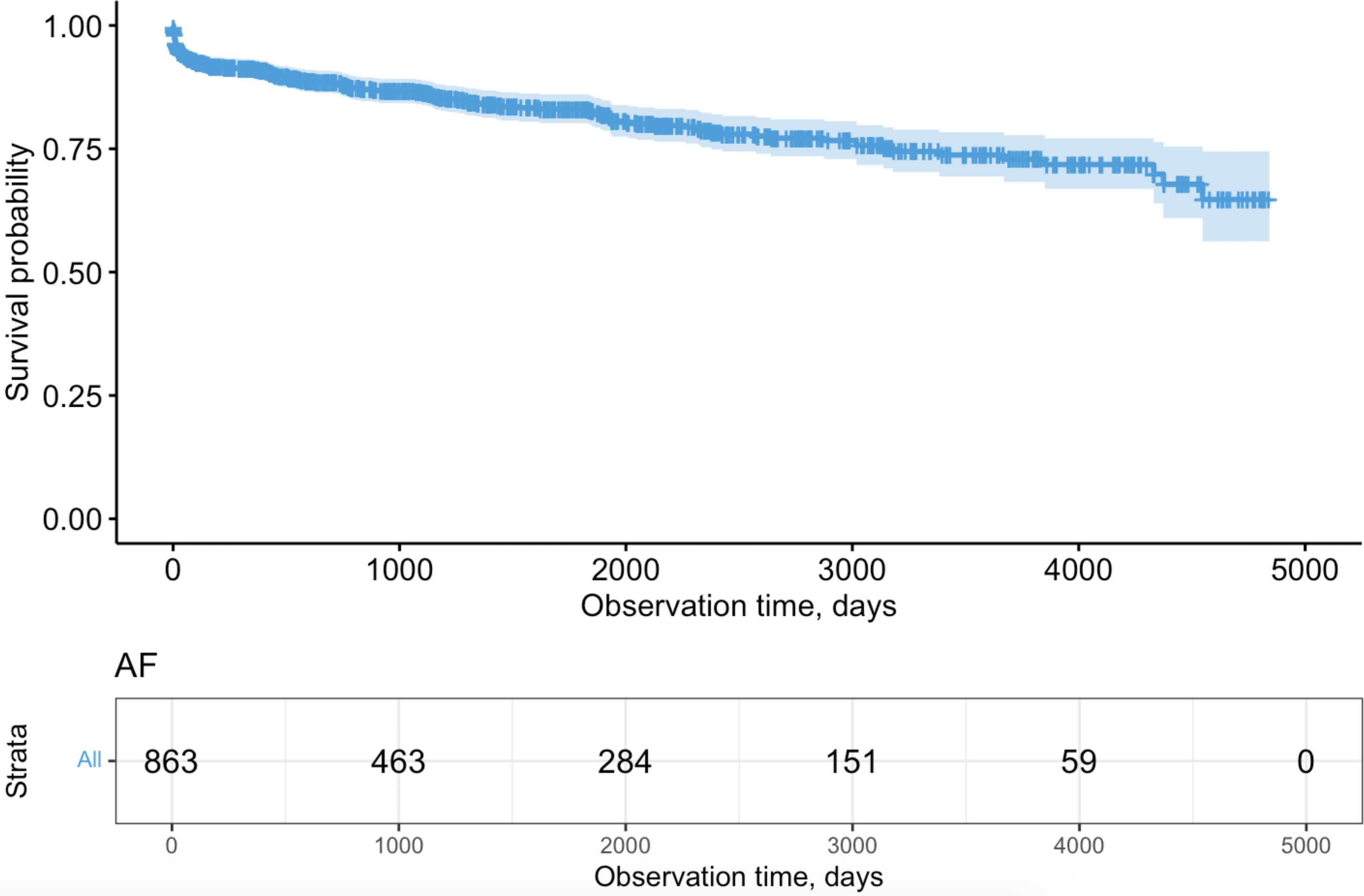
Survival curves of adverse primary and secondary outcomes in pericarditis patients.

**Figure 3.**
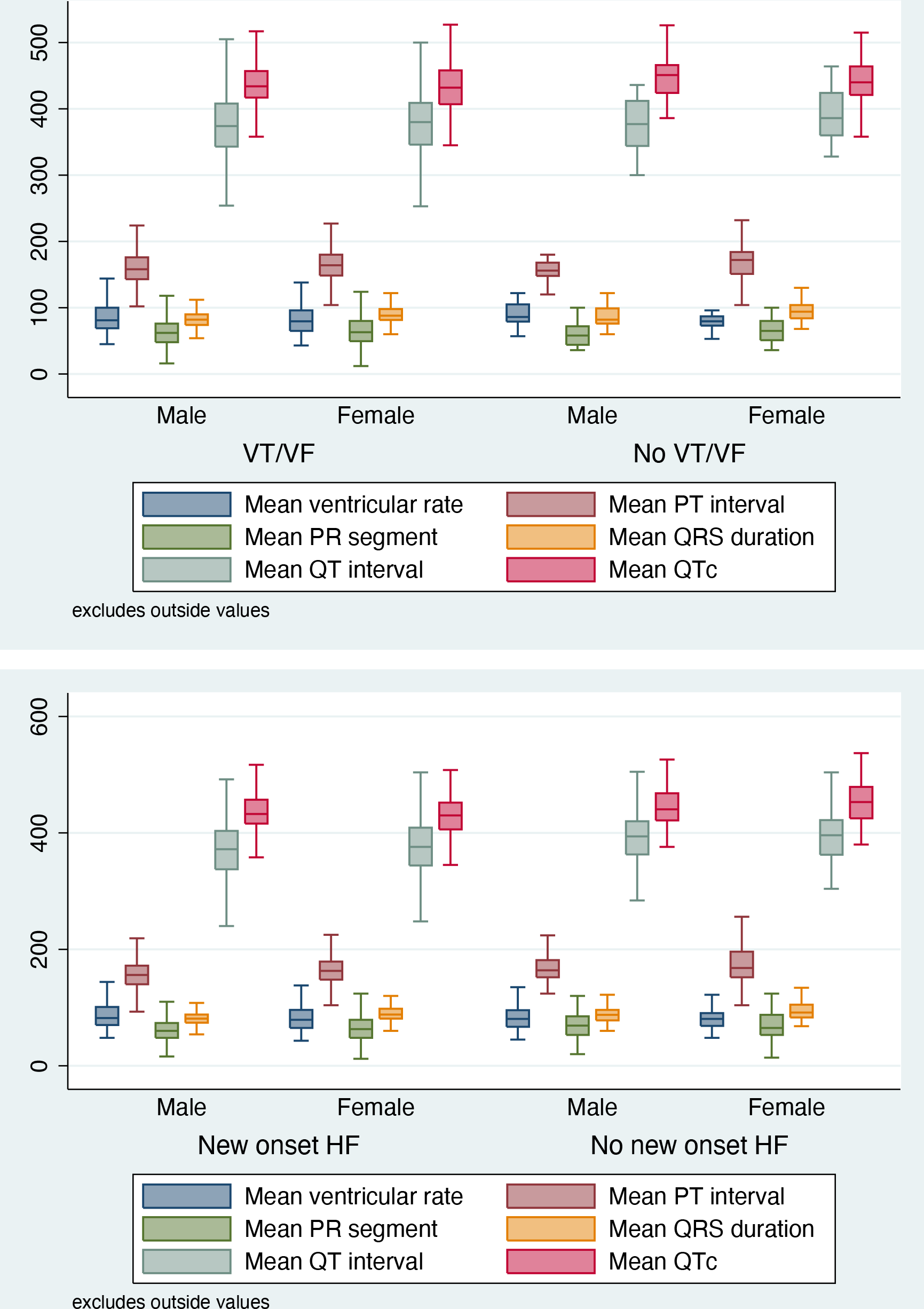
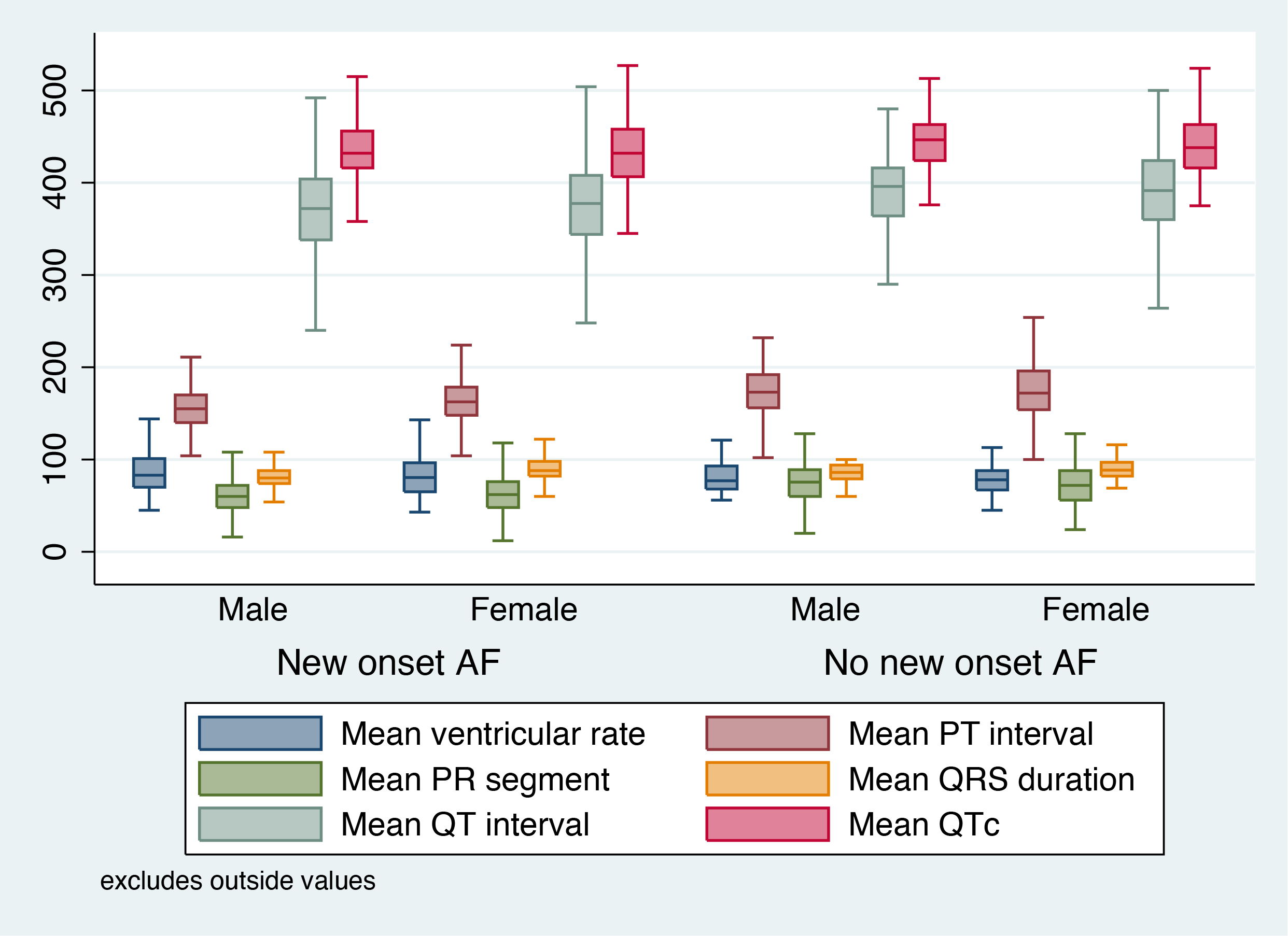
Gender-specific ECG measurements with/without VT/VF, new onset HF and new onset AF.

